# Unbiased metagenomic detection of RNA viruses for rapid identification of viral pathogens in clinical samples

**DOI:** 10.1101/2024.03.26.24304686

**Authors:** Anthony D. Kappell, Amanda N. Scholes, Matthew B. Scholz, Nicolette C. Keplinger, Leah W. Allen, Matthew C. Murray, Krista L. Ternus, F. Curtis Hewitt

## Abstract

Unbiased long read sequencing approaches for clinical metagenomic sample analysis holds enormous potential for pathogen detection, including improved detection of unknown, novel or emerging viruses. However, the rapid rate of development in nanopore sequencing and library preparation methods complicates the process of selecting a standardized method for unbiased RNA virus detection. Here, we evaluate multiple sequencing approaches to identify a workflow with sufficient sensitivity, limits of detection, and throughput for potential utilization in a clinical laboratory setting. Four separate library preparation methods for the Oxford Nanopore Technologies MinION sequencer are compared, including direct RNA, direct cDNA, rapid cDNA, and double stranded cDNA. We also establish that depletion of host RNA is not required and can be deleterious for viral RNA detection in some instances when using samples in viral transport media (VTM) or plasma. Using unbiased whole genome amplification following reverse transcription, we achieve limits of detection on the order of 1.95E03 GE/mL of Venezuelan Equine Encephalitis Virus (VEEV) spiked in human plasma. We also report initial detection of 5.43E06 GE/mL of coronavirus 229E spiked into VTM samples containing human background RNA which are expected to decrease significantly during upcoming testing. These metrics were achieved within a 6-plex multiplex reaction, illustrating the potential to increase throughput and decrease costs for relevant sample analysis. Data analysis was performed using EPI2ME Labs framework and open access tools that are readily accessible to most clinical laboratories. Taken together, this work describes an optimized method for unbiased nanopore sequencing and analysis of RNA viruses present in two common clinical matrices.

## Introduction

RNA viruses pose a significant threat to global public health. Beyond the current coronavirus pandemic, filoviruses (e.g., ebolaviruses), alphaviruses (e.g., Venezuelan Equine Encephalitis Virus (VEEV)), and flaviviruses (e.g., Zika virus), among others, cause recurring outbreaks in the Americas and around the world. While transmission pathways and replication numbers vary between them, viruses generally form the most transmissible infectious threats, and beyond vaccines, they have fewer options for treatment and containment following an outbreak. Spillover of zoonotic diseases from animal populations (e.g., bats), which is the likely cause of many recent viral outbreaks, presents a continuing threat. Unbiased, metagenomic sequence-based approaches could lead to early detection of both previously characterized and novel RNA viruses [1]. It has the potential to serve as a hypothesis-free, single, and universal assay for diagnostics of known and novel infectious disease and Emerging Infectious Diseases (EIDs) directly from samples [2–5]. Metagenomics for pathogen detection in public health could overcome many of the current challenges with traditional methods. It offers the power to identify novel or divergent pathogens for which there is no other diagnostic test available, as well as the ability to more rapidly and cost effectively identify known pathogens. Techniques that require serial testing against a list of suspected pathogens or culturing can lead to delayed actionable results especially for slow-growing pathogens, such as *Mycobacterium tuberculosis*, while metagenomic approaches comprise a single test and are increasing in speed as sequencing technologies advance. Although performing multiple tests for known pathogens can become very expensive and time consuming, the declining cost of a single metagenomic test makes it more economically justifiable. These trends of increasing speed and reduced cost are highlighted by nanopore sequencing, which can combine real time sequence analysis with relatively inexpensive, disposable sequencing reagents.

Nanopore sequencing for public health threats is well established [6–9], and targeted nanopore sequencing for viral detection has been successful as part of the COVID-19 pandemic response in public health labs [10]. Unlike unbiased metagenomic sequencing, targeted approaches selectively amplify specific sections of viral genomes before sequencing. Rapid nanopore metagenomics workflows, such as cDNA synthesis and direct RNA sequencing, have been established as a foundation for unbiased viral sequencing, but significant work is needed to evaluate and validate the best performing methods to enable the implementation of these promising new tools in public health labs. In this study, we evaluate methods using the Oxford Nanopore MinION sequencing device to detect RNA viruses rapidly and accurately in an unbiased manner. This approach capitalizes on the strengths of the sequencing device by generating sequence data for real time analysis to dramatically shorten the time required to sequence each sample, and critically, enabling workflows for unbiased sequencing to detect novel pathogens and EIDs.

## Materials and Methods

### Source Material Quantification and Contrived Sample Preparation

All experiments were performed using VEEV TC-83 (NR-63), SARS-CoV2 (NR-52286), and Human coronavirus (HCoV) 299E (NR-52726). All samples were obtained from BEI Resources. The quantity of RNA in viral stocks (source) was determined using GoTaq Probe (Promega) for VEEV and SARS-CoV2 and GoTaq RT-qPCR (Promega) for Human coronavirus (HCoV) 229E using manufacturer’s instructions with an annealing temperature of 55°C. qPCR assays utilized commercially available primers for SARS-CoV-2 (Integrated DNA Technologies Catalog #10006713) and previously established primer sets for VEEV and HCoV 229E. [11] [12] [13] VEEV TC-83 contrived sample was prepared to a working concentration of 1.0×10^11^ Genome Equivalents (GE)/mL by adding 149.15 µL VEEV TC-83 stock into 1850.85 µL K_2_EDTA human plasma (Gender Unspecified Not Filtered, 5mL (HUMANPLK2-0000285). A negative control plasma sample was prepared by adding 75.48 µL PBS to 925.42 µL human plasma. HCoV 229E contrived sample was prepared by adding 149.15 µL HCoV 229E stock into 1850.85 µL SARS-CoV-2 Swab Negative VTM, (Discovery Life Sciences) to a working concentration of 4.08E09 GE/mL. A negative control VTM sample was prepared by adding 45.70 µL PBS to 454.30 µL SARS-CoV-2 Swab Negative sample in VTM.

### RNA Extraction and host rRNA Depletion

RNA from viral stocks, control stocks, and contrived samples were extracted using the Total RNA Purification Plus Micro Kit (Norgen #48500) using manufacturer’s instructions and adapting the non-coagulated blood protocol with minor changes of input volume increased to 140 µL from 100 µL and Buffer RL increased from 350 µL to 490 µL (3.5 volumes). To deplete host rRNA, Illumina’s Ribo-Zero Plus rRNA Depletion Kit (#20040526) was used to enzymatically digest ribosomal and globin RNA, following the manufacturer’s instructions. Purified contrived RNA and depleted contrived RNA samples were quantified using GoTaq Probe (Promega) using manufacturer’s instructions using an annealing temperature of 55°C.

### Library Preparation and Sequencing

For testing which library sequencing method worked best, we directly tested Direct RNA (dRNA), Direct cDNA (DcRNA), Rapid, and Double stranded cDNA (dscDNA) sequencing methods. Direct RNA sequencing was performed by using Direct RNA sequencing kit (SQK-RNA002) with the manufacturer’s instructions. Starting RNA input of 9 µL (<500ng) was reverse transcribed using RT adapter and Superscript III (Invitrogen 18080044) and RT adapter resulting in an RNA/DNA hybrid. Direct cDNA sequencing was performed using Direct cDNA Sequencing kit (SQK-DCS109). Briefly, 7.5 µL purified RNA (<100ng) is used to generate first strand cDNA using Maxima H Minus Reverse Transcriptase (Thermofisher EP0741) using a poly T strand-switching primer. Synthesis of the second strand of cDNA occurred using 2x LongAmp Taq Master Mix (NEB M0287S) before end repair and adapter ligation. For Rapid and dscDNA sequencing, RNA (12 µL) was transcribed using Maxima H Minus Double-Stranded cDNA Synthesis Kit (Thermofisher K2561) using random hexamers and following the manufacturer’s instructions with the minor change of increasing the 1^st^ strand enzyme to 2 µL. Rapid sequencing was performed using the Rapid sequencing kit (SQK-RAD004) using the manufacturer’s instructions. dscDNA libraries were performed using the end repair and adapter ligation of either the Direct cDNA sequencing kit or Ligation sequencing kit (SQK-LSK110), following the manufacturer’s instructions. Rapid Barcode sequencing kit (SQK-RBK004) was used for multiplexing rapid libraries. Ligation sequencing kit (SQK-LSK110) and native barcode expansion kit 1-12 (EXP-NBD104) were used for multiplexing dscDNA libraries.

All bead cleanups were done on a microfuge tube magnetic separation stand (Permagen). Sequencing of libraries were performed on Oxford Nanopore MinION Mk1B or Mk1C using R9.4.1 flow cells (FLO-MIN106D) or Flongle flow cells (FLO-FLG001) with the Flongle adapter (FLGIntSP). Each flow cell was primed and loaded using manufacturer’s instructions. Each run used default settings and ran for approximately 24 hours.

### Unbiased cDNA Amplification

Addition of REPLI-g Whole Transcriptomic Analysis (WTA) Single Cell Kit (Qiagen 150063) to the dscDNA and Rapid workflows were examined for increased sensitivity. REPLI-g WTA was used for unbiased cDNA amplification with the following modifications. Briefly, for RNA samples selected for amplification, double-stranded cDNA was transcribed using random hexamers with the Maxima H Minus Double-Stranded cDNA Synthesis Kit and was cleaned using AMPure XP beads. The cleaned cDNA was then denatured at 95°C for 3 min, snap chilled on ice, and amplified using REPLI-g sc Reaction Buffer and SensiPhi DNA polymerase. Amplified samples underwent an AMPure XP bead clean up and digestion using T7 Endonuclease I (NEB M0302L). For effective removal of T7 digested fragments, a custom AMPure XP bead solution was made using PEG 8000 50%(w/v) (Fisher Scientific NC1017553). Once cleaned, library construction was performed using the dscDNA or Rapid workflows.

### Data Analysis

For each sequencing run, passing reads (default threshold of Q8) were concatenated, and quality control (QC) was performed with by removing all reads that map human before aligning to the viral reference genomes. For validation of viral sequencing and coverage, the target viral genome was selected (e.g., SARS-CoV-2 reference genome, VEEV NC_001449 reference genome, human coronavirus 229E AF304460.1 reference genome). Reads passing QC were aligned to viral genomes using minimap2 with default ont parameters. Alignments were analyzed to determine evenness, depth, and total coverage of the target genome for each library preparation method, which was intended to be used in line with the provisional guidance for sequencing-based diagnostics). Alignments to the viral genome references were used to generate a consensus sequence, BLAST was used to generated additional coverage and percent identity statistics. A presumptive match criteria threshold for evaluating workflows was set based on alignment with known reference genome sequences with greater than 90% identity over 90% or more of the genome. The threshold was determined.

## Results

### Library Preparation Method Evaluation

We first sought to characterize genome sequence coverage across four sequencing library preparation workflows. Total RNA was extracted from lysates containing SARS-CoV-2 (heat inactivated) and VEEV. The maximum nucleic acid quantity as specified by manufacturer recommendations was then used as input for each of the four library preparation workflows (Direct RNA (dRNA), Direct cDNA (DcRNA), Rapid, and Double stranded cDNA (dscDNA)). Figure 1 shows the resulting coverage across the two genomes for each library preparation method. (Additional statistics are available in Table S1) The different sequence library preparation workflows produced different levels of coverage of the viral genomes. Because the DcDNA and dRNA methods require the use of a poly-dT primer for reverse transcription, these methods produced a bias for coverage at the 3’ end of the genome due to the reliance on the poly-A tail for primer annealing and ligation, respectively. The dscDNA and Rapid method both utilized a random-hexamer for reverse transcription, reducing the bias of specific genomic regions and allowing sequencing of any RNA viruses, not just those with a poly-A tail. As a result, the dscDNA and Rapid methods performed better overall in terms of total genome coverage.

**Figure 1.**
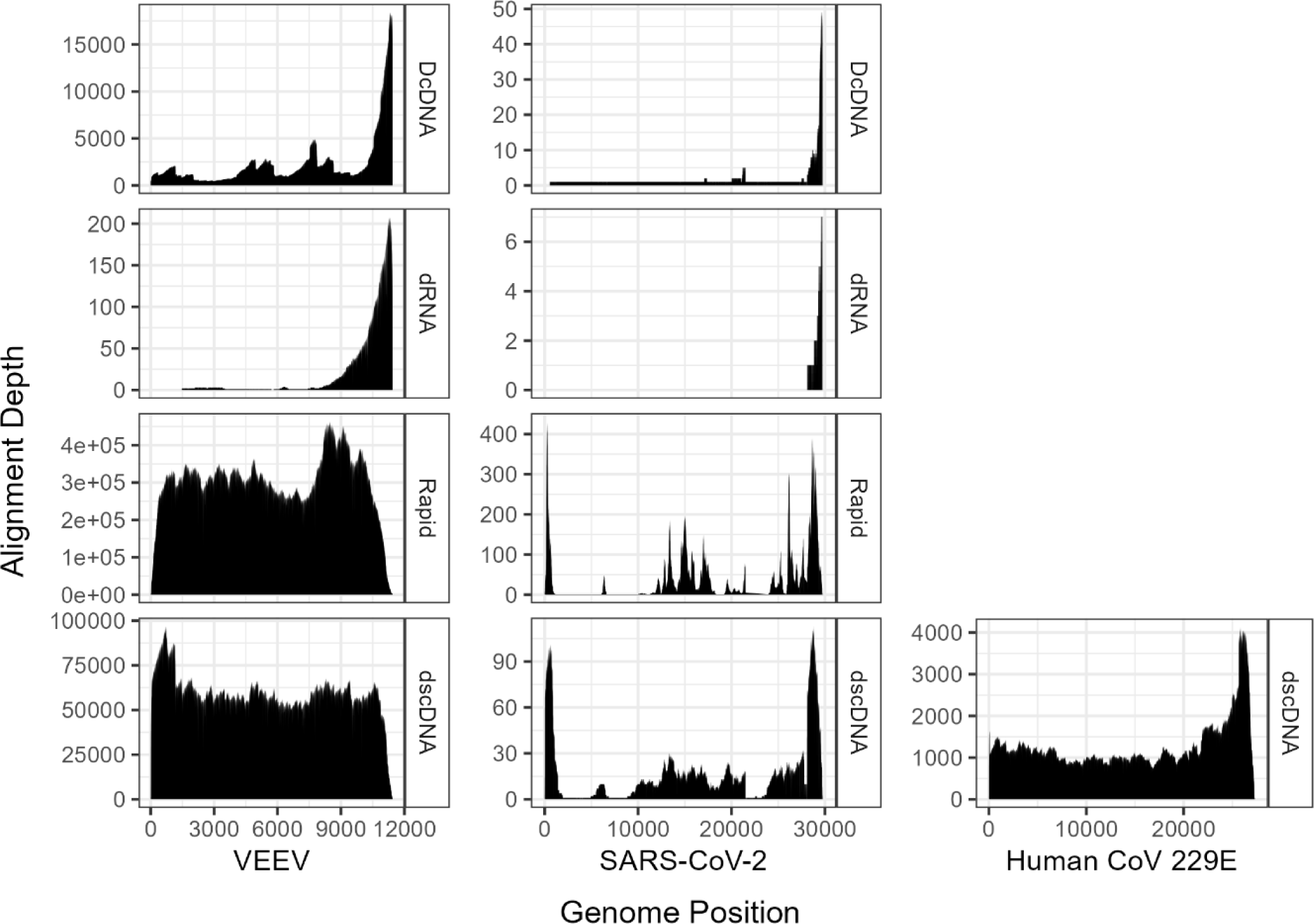
Coverage of the viral RNA genomes based on depth of read alignment.

When comparing genome sequence coverage between VEEV and SARS-CoV-2, it was apparent that the coronavirus material had lower sequence coverage overall. This discrepancy in read counts could be attributed to lower starting input (∼4.23E7 GE SARS-CoV-2 vs. ∼8.27E9 GE VEEV). The SARS-CoV-2 material had a higher host background than VEEV based on the abundance of reads mapping to the viral vs. cell culture component of the RNA extracts (data not shown). We hypothesized that, because the SARS-CoV-2 material was heat inactivated, there may have been substantial genome fragmentation and degradation of the SARS-CoV-2 genomes, resulting in lower overall read counts and percentage of passing quality reads. To test this, an alternate, non-inactivated coronavirus culture, Human coronavirus 229E (HCoV 229E), was examined in parallel. Following library preparation using the dscDNA method, it was apparent that HCoV 229E showed markedly higher genome sequence coverage (Figure 1), as well as higher read count and percentage of passing quality reads compared to SARS-CoV-2 (Table S1). This was the case even as HCoV 229E had a similar percentage of reads mapped to the viral genome as SARS-CoV-2 in the respective sequencing runs. Despite the low quality of RNA derived from the heat inactivated SARS-CoV-2 material, the sequencing results demonstrated clear benefits of the random hexamer based reverse transcription methods of dscDNA and Rapid. The Rapid and dscDNA method had greater coverage over the genome compared to DcDNA and dRNA. The dRNA methods using VEEV source material produced low coverage of the genome, while the DcDNA method produced a mean sequencing depth orders of magnitude lower than the Rapid or dscDNA methods using the VEEV source material.

Next, we sought to evaluate performance across a number of critical metrics for the performance of metagenomic sequencing assays across the DcDNA, dRNA, dscDNA, and Rapid sequencing workflows. To do this, contrived samples were assembled to mimic human clinical samples. VEEV source material was spiked into human plasma prior to RNA extraction and HCoV 229E was spiked into remnant clinical viral transport media (VTM) samples. Samples were then extracted and sequenced. VEEV samples were sequenced by all four sequencing workflows, while HCoV 229E samples were only sequenced using the top two performing workflows (Rapid and dscDNA) to conserve sequencing reagents.

The dscDNA and Rapid methods yield a greater number of reads mapped to VEEV compared to DcDNA and dRNA (Figure 2). dRNA had a greater total number of reads compared to DcDNA and dscDNA but less mapped VEEV reads. The dcDNA method produced the lowest read count of all library preparation methods. The dscDNA and Rapid methods also generated higher quality data suitable for use in mapping and database alignments (Figure 3). Similar to the direct sequencing of VEEV viral extract, the dRNA method resulted in low mean read depth of the genome from RNA extracted from active VEED spiked in human plasma. The DcDNA method using VEEV spiked plasma also had relatively low mean read compared to dscDNA and Rapid methods.

**Figure 2.**
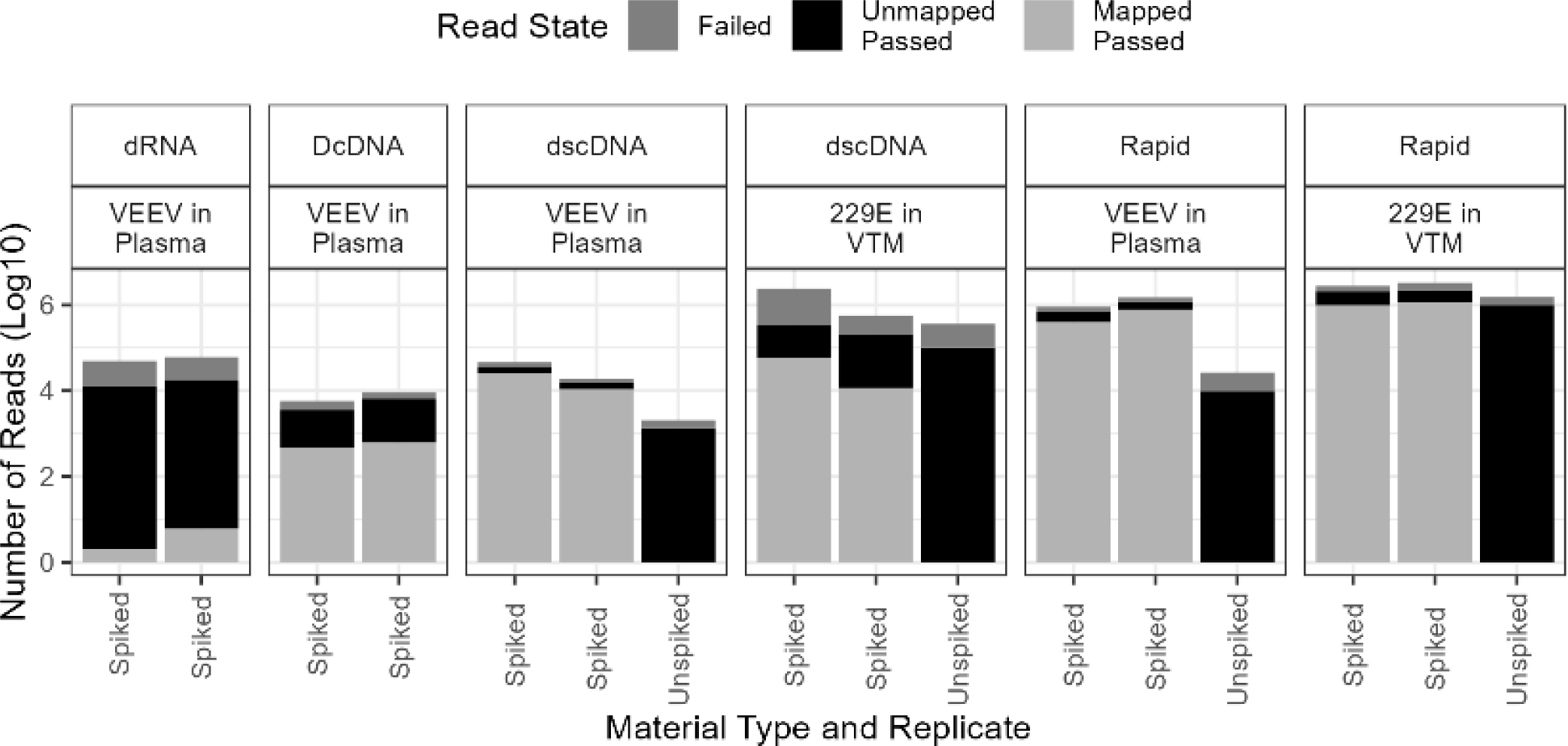
Passed and failed reads states of the difference sequencing workflows for contrived viral samples.

**Figure 3.**
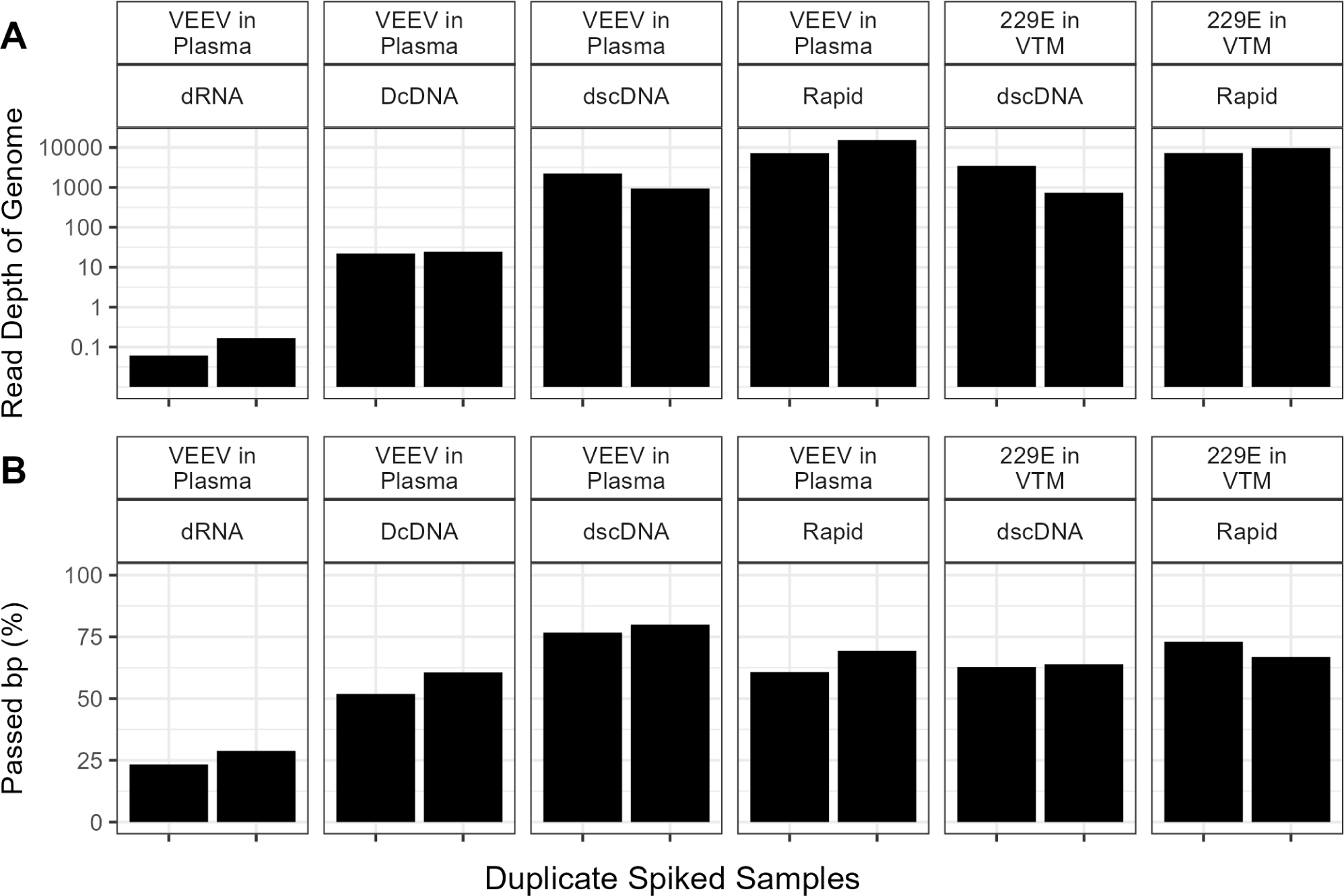
Average read depth (left) and percent passed bp data (right) from the different sequencing workflows.

With clear performance benefits apparent for the dscDNA and Rapid methods, we utilized these methods to evaluate HCoV 229E detection in a VTM sample background. HCoV 229E was sequenced and detected in a similarly robust manner compared to VEEV in terms of read depth (Figure 2) and read quality and assessment (Figure 3). Summary metrics for both VEEV and HCoV 229E spiked samples are shown in Table 1 for the dscDNA and Rapid methods, including non-spiked control samples comprised of RNA extracted from plasma or VTM not containing virus. (Additional metrics are available in Table S2). The table contains the relative genomic equivalence of viral genomes used in each sample input to workflows based on RT-qPCR within the contrived sample indicating the viral load. This value may be an overestimate of certain viral genomic copies, especially for HCoV 229E wherein the RT-qPCR target of the N gene falls within a sub-genomic transcript that may be in greater abundance than the whole genome in the viral lysates used for this study. This sub-genomic transcript at the 3’ end of the genome maybe noticeable in Figure 1.

**Table 1.** Sequencing Summary of Contrived Viral Samples Using dscDNA or Rapid Method.

| Method | dscDNA | dscDNA | dscDNA | dscDNA | dscDNA | dscDNA | Rapid | Rapid | Rapid | Rapid | Rapid | Rapid |
| --- | --- | --- | --- | --- | --- | --- | --- | --- | --- | --- | --- | --- |
| Virus | VEEV | VEEV | 229E | 229E | None | None | VEEV | VEEV | 229E | 229E | None | None |
| Matrices | Plasma | Plasma | VTM | VTM | Plasma | VTM | Plasma | Plasma | VTM | VTM | Plasma | VTM |
| Replicate | 1 | 2 | 1 | 2 | 1 | 1 | 1 | 2 | 1 | 2 | 1 | 1 |
| GE in total | 1.04E+08 | 1.04E+08 | 1.33E+09 | 1.33E+09 | 0 | 0 | 1.04E+08 | 1.04E+08 | 1.33E+09 | 1.33E+09 | 0 | 0 |
| Mean Depth | 937.10 | 2236.30 | 734.54 | 3461.83 | 0 | 0 | 15373.00 | 7265.00 | 9688.86 | 7301.84 | 0 | 0 |
| Coverage (%<br>minimap2) | 99.97 | 99.99 | 100.00 | 100.00 | 0 | 0 | 99.99 | 99.99 | 100.00 | 100.00 | 0 | 0 |
| Identity of<br>Coverage (%) | 96.34 | 96.33 | 99.91 | 99.91 | 0 | 0 | 96.36 | 96.35 | 99.93 | 99.93 | 0 | 0 |
| N50 | 1,209 | 1,221 | 1,535 | 1,962 | 658 | 1317 | 303 | 280 | 349 | 331 | 217 | 261 |
| Longest<br>Alignment<br>length (Kb) | 4.62 | 4.46 | 6.11 | 6.87 | 0 | 0 | 2.07 | 2.08 | 3.14 | 2.61 | 0 | 0 |
| Total Reads | 19,094 | 46,177 | 575,045 | 2,411,913 | 2047 | 361,490 | 1,498,299 | 911,560 | 3,313,337 | 2,788,740 | 26,785 | 1,536,316 |
| Passing<br>Quality Reads<br>(%) | 80.22 | 77.97 | 35.15 | 13.94 | 65.5 | 28.07 | 78.84 | 75.00 | 66.71 | 74.25 | 35.89 | 65.03 |
| Mapped<br>Passed Reads<br>to Virus (%) | 69.92 | 73.00 | 5.68 | 17.04 | 0 | 0 | 67.08 | 57.42 | 52.22 | 47.42 | 0 | 0 |
| Passing bp<br>(%) | 79.99 | 76.73 | 63.75 | 62.63 | 65.5 | 28.07 | 69.36 | 60.75 | 66.88 | 73.04 | 35.89 | 60.26 |

### Assessment of Need for Host RNA Depletion from Contrived Samples

Depletion of host RNA within metagenomic samples is commonly used to enrich for viral and other microbial signatures. We examined the effect of host RNA depletion on contrived VEEV samples within a human plasma background. Table 2 summarizes the results from sequencing of RNA recovered following depletion of human ribosomal RNA (cytoplasmic and mitochondrial) and globin RNA (Additional metrics available in Table S3). This included replicate sequencing of the contrived samples using both dscDNA and Rapid library prep methods.

**Table 2.**
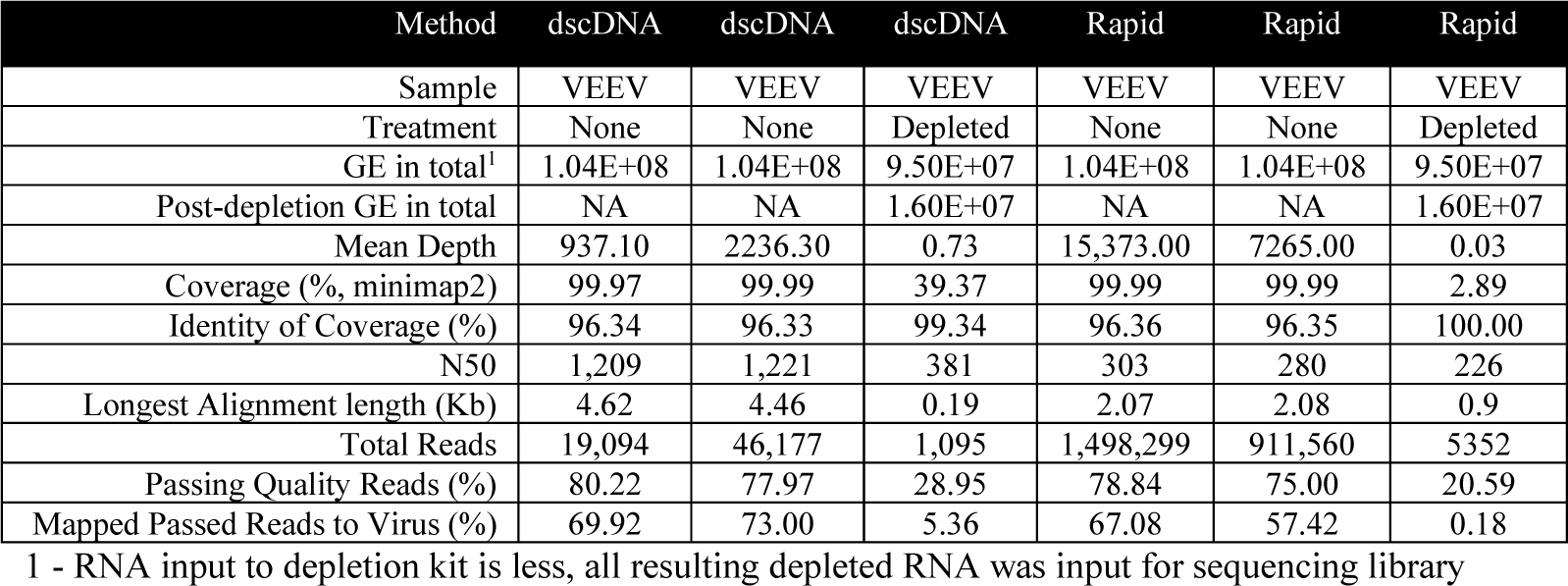
Sequencing Summary of Host Depletion from Contrived Viral Samples.

In all cases, the host depletion method used caused a decrease in total number of human reads as expected, but the percent of reads passing quality filters and mapping to the viral genome also decreased significantly. This may be due to the method used by the depletion kit, wherein target RNA is enzymatically digested following hybridization by DNA probes complementary to the target sequence. Based on the N50 and the decrease in longest read within dscDNA method, the enzymatic digest likely targeted non-specifically to the total RNA. This non-specific cleavage could be related to the low total RNA input into the reaction or the relatively low abundance of human RNA in the samples. This method and alternative methods for depletion may be necessary for other sample types with high host background (e.g., whole blood), but the relatively low abundance of human RNA within the plasma and VTM samples does not appear to necessitate host RNA depletion.

We utilized centrifuge to assign taxonomical classification to the sequenced reads (Figure 4). There were 2-to 3-log decreases in the VEEV genomic material recovered from the samples that were depleted compared to non-depleted. Meanwhile, human RNA was only depleted by 1-to 2-log. There was very little reduction of human RNA depleted within the plasma sample itself.

**Figure 4.**
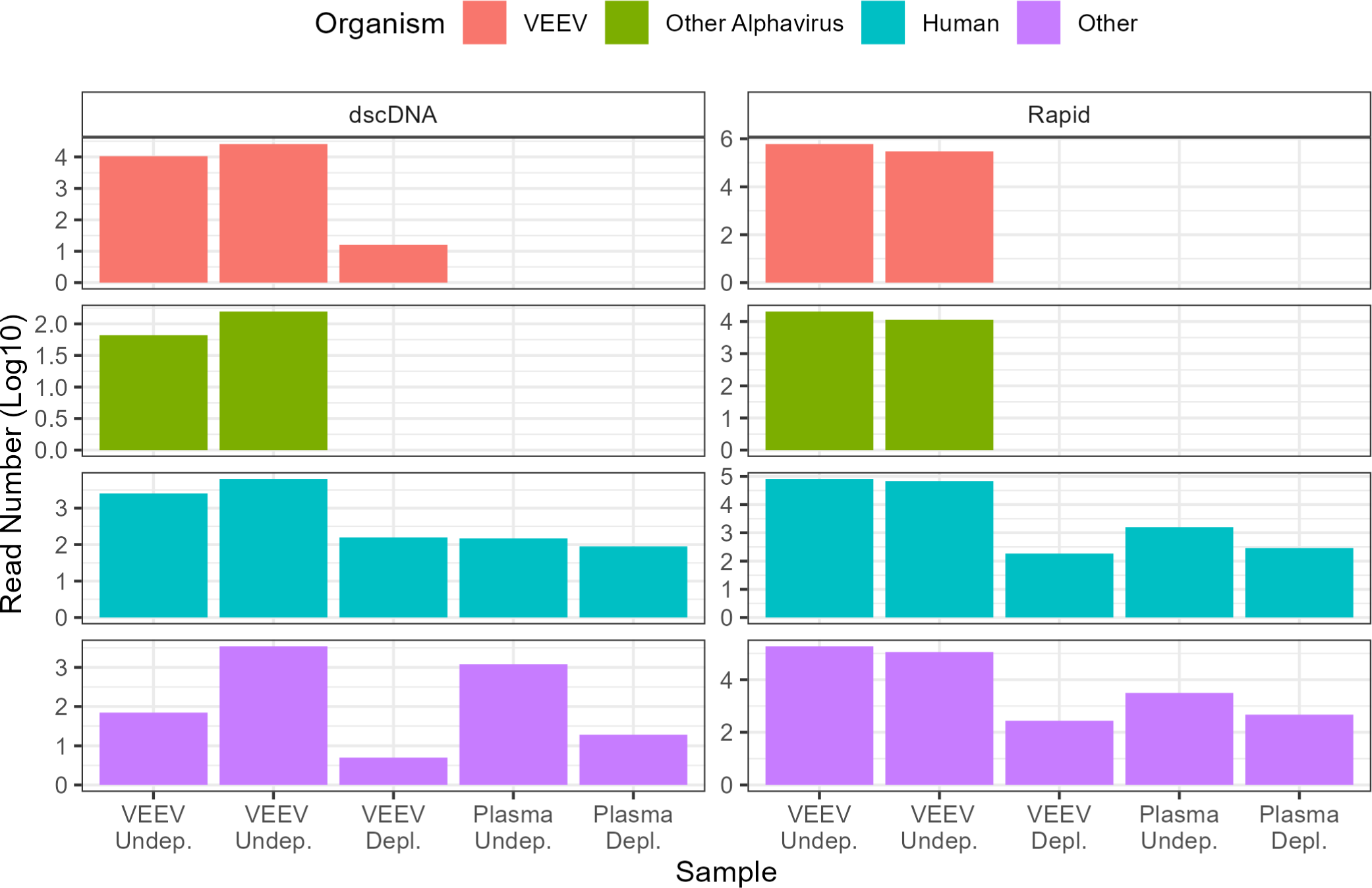
Effect of Depletion on Centrifuge Read Counts to Different Taxonomy.

### Assessment of Sample Multiplexing by Barcoding

There are few approaches for sample multiplexing to reduce per sample sequencing costs. Barcoding for running multiple samples on a single flow cell can reduce time by allowing parallel sample preparations and cost by reducing the number of flow cells needed to process multiple samples. However, multiplexing can also reduce sensitivity by producing fewer sequencing reads per samples or barcode and introduce the potential for cross contamination of sample read pools because of barcode crosstalk. The Rapid method essentially allows the barcodes to be added during tagmentation while the dscDNA method allows the use of the Native Barcoding Kit to ligate indexes to each cDNA molecule.

We used the 12-plex based barcoding kits to examine multiplexing of the dscDNA and Rapid methods. Ten of the samples contained duplicate reactions of RNA extracted from human plasma spiked with VEEV at multiple concentrations. The last two samples contained RNA extracted from A549 cells and human plasma. Tables 3 and 4 contain the summary results of sequencing using a 12-plex dscDNA method and Rapid method, respectively.

**Table 3.**
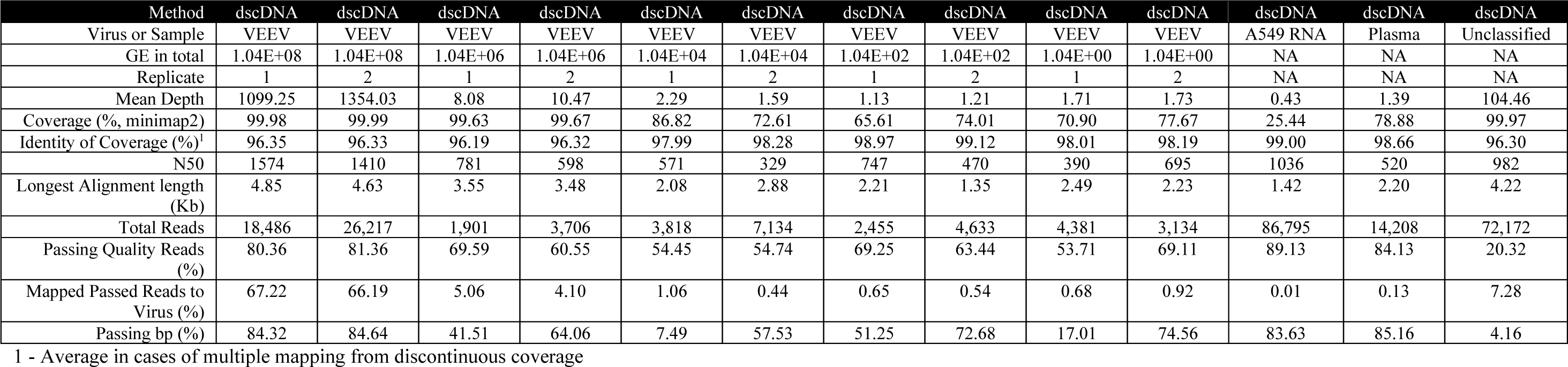
Sequencing Summary of Multiplex (12 samples) of dscDNA Method.

**Table 4.**
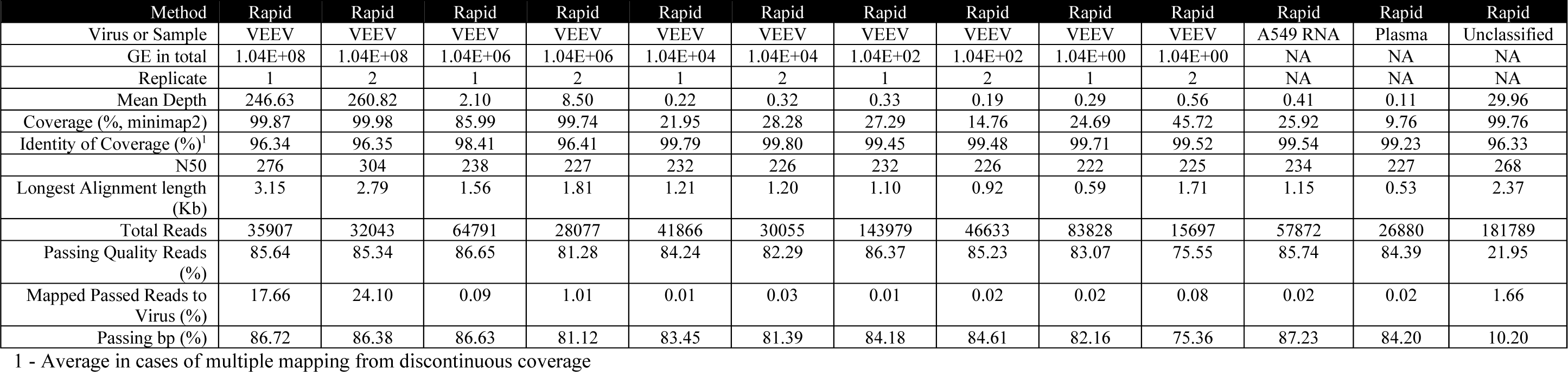
Sequencing Summary of Multiplex (12 samples) of Rapid Method.

The sensitivity threshold for positive detection was set at > 90% identify over > 90% of the genomic sequence. The multiplex dscDNA method (Table 3) dropped below this threshold with VEEV load at 1.04E04 GE of VEEV in plasma with coverages at 88% and 75% for the duplicate library preparations. The multiplex Rapid method (Table 4) showed a similar drop in coverage at this loading but to a greater extent (24% and 34%). (Additional metrics are available in Table S4 and S5 for multiplex dscDNA and Rapid methods, respectively)

The total read counts produced by the multiplex dscDNA method at 1.04E8 load was comparable to the single-plex of the method at the same loading. In addition, the percent of passing reads, percentage of reads mapped to VEEV, and the mean depth were also comparable. Upon the analysis of the duty-time record for the multiplex and single-plex of dscDNA method (Figure 5), single-plex utilization of the available pores appeared to be low. This low utilization provided additional pores for sequencing for multiplex of additional dscDNA libraries without decreasing read quality and read number.

**Figure 5.**
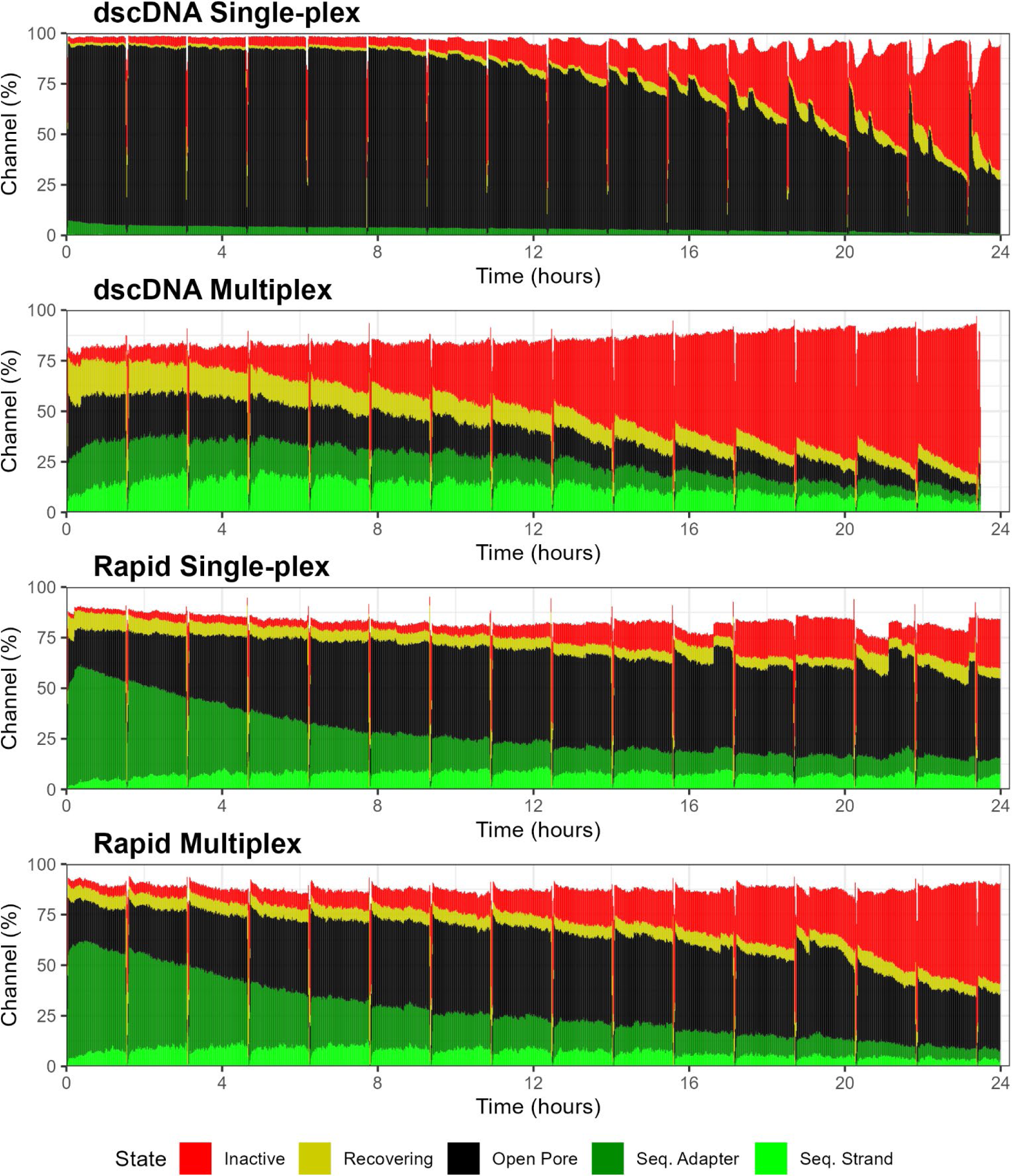
Duty time record of dscDNA and Rapid Single-plex and Multiplex (Pore-Scan)

The total number of reads using the Rapid method decreased from millions of reads in the single-plex workflow to tens-of-thousands of reads in the multiplex at the same loading. The percentage of quality passing reads was higher in the multiplex compared to the single-plex, but percentage of mapped reads was lower in the multiplex leading to a reduction in mean depth. Some data was lost due to the inability to assign to a barcode (unclassified). Analysis of the duty-time record for the multiplex and single-plex Rapid method (Figure 5) had comparable percentage of pores actively sequencing, suggesting that the Rapid method in single-plex or multiplex used similar flow cell capacity available for sequencing.

### Assessment of Lower Throughput Flongle

We evaluated the flongle flow cell. The flongle has lower throughput and cost compared to the standard flow cell. We evaluated the dscDNA and Rapid method for use on the flongle using RNA extracted from plasma spiked with VEEV. Table 5 shows a summary of sequence results from using flongle in place of a standard flow cell for the dscDNA and Rapid methods. Total read counts were significantly lower for dscDNA and Rapid methods using the flongle compared to the standard flow cell. The dscDNA method had comparable percentage of passing quality reads and mapped reads using the flongle to that of a standard flow cell. The Rapid method using the flongle had a decrease in percentage of passing quality of the reads and mapped reads to that of the standard flow cell.

**Table 5.** Sequencing Summary of Flongle Use.

| Method | dscDNA | Rapid |
| --- | --- | --- |
| Virus | VEEV | VEEV |
| GE in total | 1.04E+08 | 1.04E+08 |
| Mean Depth | 619.94 | 1.22 |
| Coverage (% minimap2) | 99.99 | 70.37 |
| Identity of Coverage (%) | 96.32 | 99.15 |
| N50 | 1434 | 208 |
| Longest Alignment length (Kb) | 4189 | 496 |
| Total Reads | 16724 | 4285 |
| Passing Quality Reads (%) | 57.21 | 20.65 |
| Mapped Passed Reads to Virus (%) | 66.71 | 9.27 |
| Passing bp (%) | 54.30 | 4.23 |

Both methods showed high inactivity of pores on the flongle with high percentage of usage of available pores based on the duty-time report (Figure 7). Interestingly, the low usage of the pores on the standard flow cell by dscDNA suggest that the flongle should provide sufficient sequence depth to align with the standard flow cell in these samples. The utilization of number of pores on the flongle was approximately 32 while the standard flow cell showed early usage at approximately 64 and falling quickly to 32. However, the increasing inactivation of pores on the flongle quickly reduced the capacity for both methods and causing low read counts. This high inactivation rate reduced the comparable output of the flongle to the standard flow cell for the dscDNA method. Future iterations of the flongle or luck, as loading often is the cause of pore inactivity, may make the flongle a future option for low input samples such as plasma or VTM.

**Figure 6.**
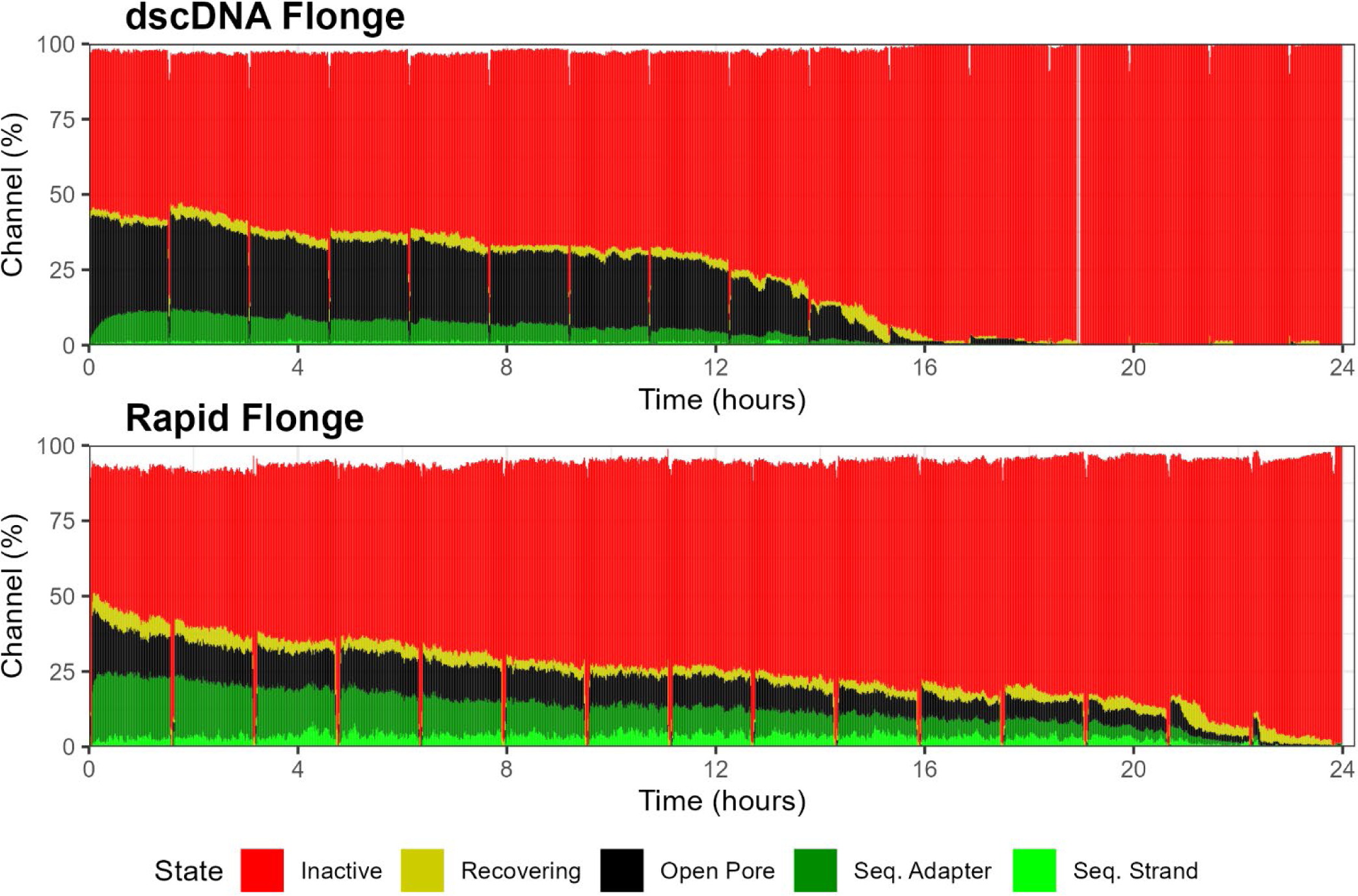
Duty Time report from dscDNA and Rapid loaded on a flongle.

**Figure 7.**
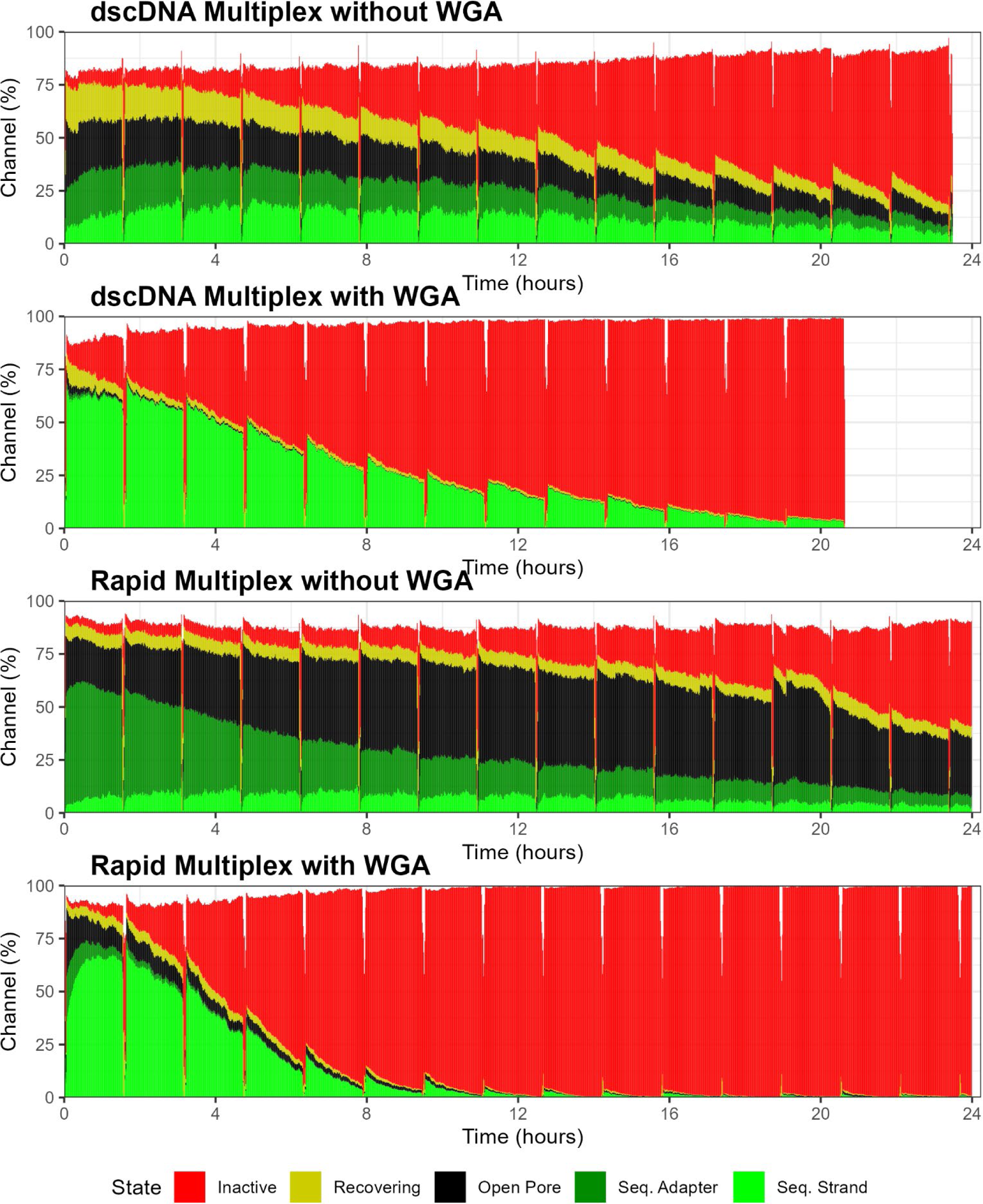
dscDNA or Rapid multiplex with and without WGA.

### Assessment of Whole Transcriptome Amplification

Because of the low RNA yields contrived VEEV samples in VTM and plasma, we evaluated whole transcriptome amplification (WTA). WTA to increase sequencing material would allow for full utilization of the available pores within a standard flow cell. WTA was examined using a 6-plex barcoding scheme using the native barcoding kit for dscDNA and rapid barcoding kit instead of rapid sequencing kit for the Rapid method. The six samples contained duplicate reactions of RNA extracted from Human plasma spiked with VEEV at decreasing viral load. The genomic equivalency entering the WTA was 8.21E05, E03, and E01.

The use of WTA had impacts on both methods. Table 6 and Table 7 contain the sequencing summary for the dscDNA and Rapid methods supplemented with WTA using VEEV, respectively. (Additional metrics are available in Tables S7 and S8). The dscDNA and Rapid method both had increases in total read count compared to the previous multiplex run. WTA greatly increased total read counts for dscDNA compared to single-plex while Rapid increase of total read counts was less, most likely due to their respective pore usages compared to the previous multiplex and single-plex runs. The percentage of passed reads and mapped reads to virus were comparable to previous multiplex runs. There was an increase in N50 in both Rapid and dscDNA methods when using WTA suggesting potentially longer reads. Both methods also had increased coverage of the genome and mean depth at lower VEEV loading. The Rapid method utilizing WTA met threshold (>90% identify over >90% of the genome) as low as 8.21E03 GE improving upon the drop at 1.04E04 GE on the previous multiplex run without WTA. The dscDNA met threshold for all viral loads tested as low as 8.21E01 GE improving greatly upon the previous multiplex run drop of sensitivity at 1.04E04 GE. Table 8 contains the sequencing summary for dscDNA method supplemented with WTA using HCoV 229E. HCoV 229E was tested as low as 2.29E05 GE viral loading resulting in positive threshold values for identification.

**Table 6.** Sequencing Summary of Whole Genome Amplification and dscDNA Method using VEEV (Multiplex of 6 samples)

| Method | dscDNA | dscDNA | dscDNA | dscDNA | dscDNA | dscDNA |
| --- | --- | --- | --- | --- | --- | --- |
| Virus or Sample | VEEV | VEEV | VEEV | VEEV | VEEV | VEEV |
| GE in total | 8.21E+05 | 8.21E+05 | 8.21E+03 | 8.21E+03 | 8.21E+01 | 8.21E+01 |
| Replicate | 1 | 2 | 1 | 2 | 1 | 2 |
| Mean Depth | 2954.70 | 3914.28 | 150.02 | 137.55 | 47.41 | 66.24 |
| Coverage (% minimap2) | 99.99 | 99.97 | 98.34 | 98.44 | 90.33 | 92.42 |
| Identity of Coverage (%) | 96.34 | 96.37 | 96.29 | 96.33 | 96.94 | 97.70 |
| N50 | 3376 | 2940 | 2734 | 3476 | 3483 | 3496 |
| Longest Alignment length (Kb) | 3.19 | 2.96 | 1.84 | 3.15 | 1.84 | 2.86 |
| Total Reads for a Barcode | 98649 | 139714 | 136040 | 108623 | 82638 | 120026 |
| Percent of Total Reads in Run | 10.69 | 15.14 | 14.74 | 11.77 | 8.96 | 13.01 |
| Passing Quality Reads (%) | 68.51 | 68.40 | 69.74 | 68.21 | 71.28 | 68.91 |
| Mapped Passed Reads to Virus (%) | 25.89 | 26.83 | 0.96 | 1.01 | 0.48 | 0.51 |
| Passing bp (%) | 66.86 | 66.20 | 67.02 | 65.28 | 68.15 | 66.83 |

**Table 7.** Sequencing Summary of Whole Genome Amplification and Rapid Method using VEEV (Multiplex of 6 samples)

| Method | Rapid | Rapid | Rapid | Rapid | Rapid | Rapid |
| --- | --- | --- | --- | --- | --- | --- |
| Virus or Sample | VEEV | VEEV | VEEV | VEEV | VEEV | VEEV |
| GE in total | 8.21E+05 | 8.21E+05 | 8.21E+03 | 8.21E+03 | 8.21E+01 | 8.21E+01 |
| Replicate | 1 | 2 | 1 | 2 | 1 | 2 |
| Mean Depth | 5056.20 | 5176.32 | 100.84 | 114.58 | 5.31 | 3.62 |
| Coverage (% minimap2) | 99.59 | 99.27 | 97.91 | 97.98 | 62.48 | 71.49 |
| Identity of Coverage (%)<sup>1</sup> | 96.34 | 96.34 | 96.35 | 96.25 | 96.97 | 96.82 |
| N50 | 2578 | 2658 | 2572 | 2661 | 2464 | 2529 |
| Longest Alignment length (Kb) | 2.71 | 2.62 | 2.10 | 1.93 | 1.75 | 1.89 |
| Total Reads for a Barcode | 188566 | 185313 | 156451 | 162121 | 213392 | 133602 |
| Percent of Total Reads in Run | 15.03 | 14.77 | 12.47 | 12.92 | 17.01 | 10.65 |
| Passing Quality Reads (%) | 90.06 | 89.37 | 89.74 | 89.05 | 89.90 | 89.93 |
| Mapped Passed Reads to Virus (%) | 25.43 | 25.98 | 0.58 | 0.60 | 0.02 | 0.03 |
| Passing bp (%) | 88.52 | 88.10 | 88.27 | 87.40 | 88.93 | 89.20 |

1 - Average in cases of multiple mapping from discontinuous coverage

**Table 8.** Sequencing Summary of Whole Genome Amplification and dscDNA Method using HCoV 229E (Multiplex with 6 samples)

| Method | dscDNA | dscDNA | dscDNA | dscDNA | dscDNA | dscDNA |
| --- | --- | --- | --- | --- | --- | --- |
| Virus or Sample | 229E | 229E | 229E | 229E | 229E | 229E |
| GE in total | 2.29E+07 | 2.29E+07 | 2.29E+06 | 2.29E+06 | 2.29E+05 | 2.29E+05 |
| Replicate | 1 | 2 | 1 | 2 | 1 | 2 |
| Mean Depth | 3566.62 | 3050.51 | 1572.42 | 1562.44 | 2059.74 | 1612.87 |
| Coverage (% minimap2) | 99.98 | 100 | 99.94 | 99.92 | 99.86 | 99.95 |
| Identity of Coverage (%) | 99.89 | 99.91 | 99.91 | 99.92 | 99.89 | 99.89 |
| N50 | 3164 | 3867 | 3434 | 3255 | 3305 | 3097 |
| Longest Alignment length (Kb) | 3419 | 3193 | 3216 | 314 | 2822 | 2340 |
| Total Reads for a Barcode | 159507 | 134289 | 150470 | 152969 | 175653 | 135563 |
| Percent of Total Reads in Run | 13.10 | 11.03 | 12.36 | 12.56 | 14.42 | 11.13 |
| Passing Quality Reads (%) | 83.53 | 78.13 | 80.41 | 81.00 | 79.07 | 80.52 |
| Mapped Passed Reads to Virus (%) | 36.50 | 34.49 | 18.15 | 18.32 | 23.24 | 23.60 |
| Passing bp (%) | 83.03 | 81.77 | 80.30 | 80.22 | 78.02 | 79.37 |

Duty-time reports (Figure 8) indicate a high rate of inactivation of pores using the WTA in both dscDNA and Rapid compared to multiplex and single-plex methods without WTA. Both methods showed initial burst of sequencing and utilization of all available pores until inactive channels disrupted productive sequencing. This inactivity was more pronounced in the Rapid method compared to the dscDNA methods, which also contributed to lower total read counts for the Rapid method. The relatively high activity very early with the dscDNA method combined with WTA explains the much higher read count and improvement of coverage and mean depth in samples with lower concentration of spiked VEEV into Human plasma. Interestingly, the barcode crosstalk was lower compared to the previous multiplex run (See Table S7-S9 for unused and unclassified barcodes). This may be due to higher concentration of starting material input to the Rapid Barcoding Kit and the Native Barcoding with Ligation Sequencing Kit. The improved sensitivity and potential for multiplexing samples (at least 6-plex) makes the use of WTA in conjunction with the Rapid and dscDNA methods superior to the methods without WTA.

**Figure 8.**
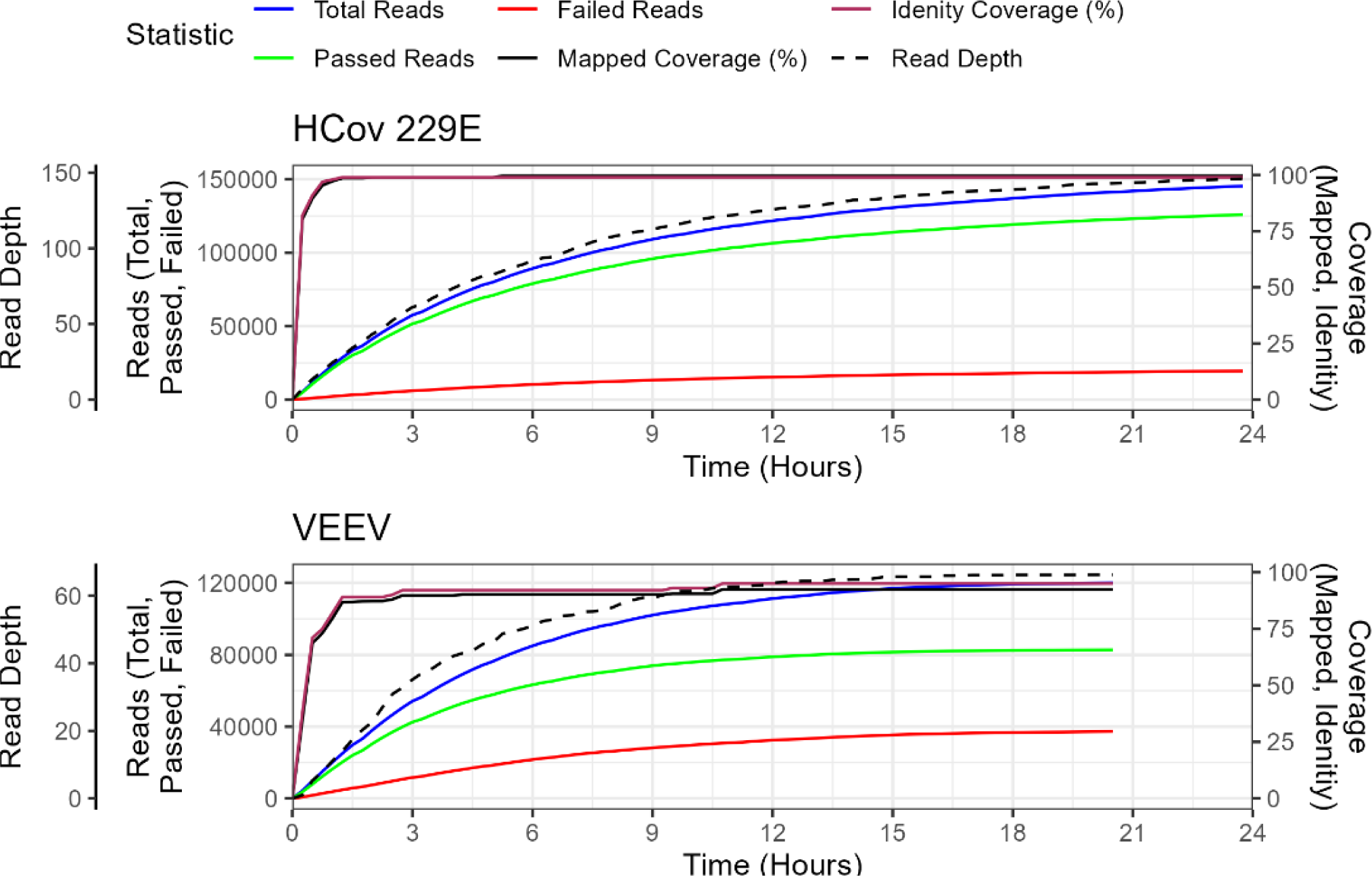
Whole Genome Amplification and dscDNA Method with HCoV 229E and VEEV indicating the Coverage and Identity of Mapped Reads Generated During the Run.

### Analysis of Clinical Remnant Samples

Clinical remnant samples consisting of respiratory swabbed samples stored in viral transport medium were subjected to the workflow from viral RNA extraction, library preparation, and bioinformatic workflow. The clinical samples did not consist of the viruses tested for the validation but are relevant RNA viruses. The majority of detected viral sequences did not rise to the set threshold of ≥90% identity over ≥90% of the genome, nor the minimum working mean depth. The only exception was the detection of human respiratory syncytial virus wildtype strain B1 (RSV) in one of the six multiplexed samples, at 96.25% identity with 99.32% coverage of the genome (Table 9). (Additional information is available in Table S10 and S11). This sample also showed 248.9 mean depth, greater than the working cutoffs of 10 or 15. While RSV was also detected in the other samples, the highest mean depth was 3.09, well below the mean depth cutoff of 10 utilized to avoid false positives due to barcode cross talk, as well as the alternative cutoffs determined be the negative control. This result also highlights the importance of negative controls for the evaluation of potential false positives and appropriate thresholds. Interestingly, after removal of RSV from the results, barcode17 had human metapneumovirus as the next top hit in both coverage (29.04%) and mean depth (0.673), including a percent identity of 92.20% of the covered genome. The detection of human metapneumovirus is the original detection using current methods. Barcode 21, after removal of RSV, has a Rhinovirus detection with two genomes at coverage of 17.58% and 16.05% with low mean depth at 0.23 and 6.71, respectively. HRV89 was detected with higher mean depth but marginally lower coverage percent identity of 79.79%.

**Table 9.**
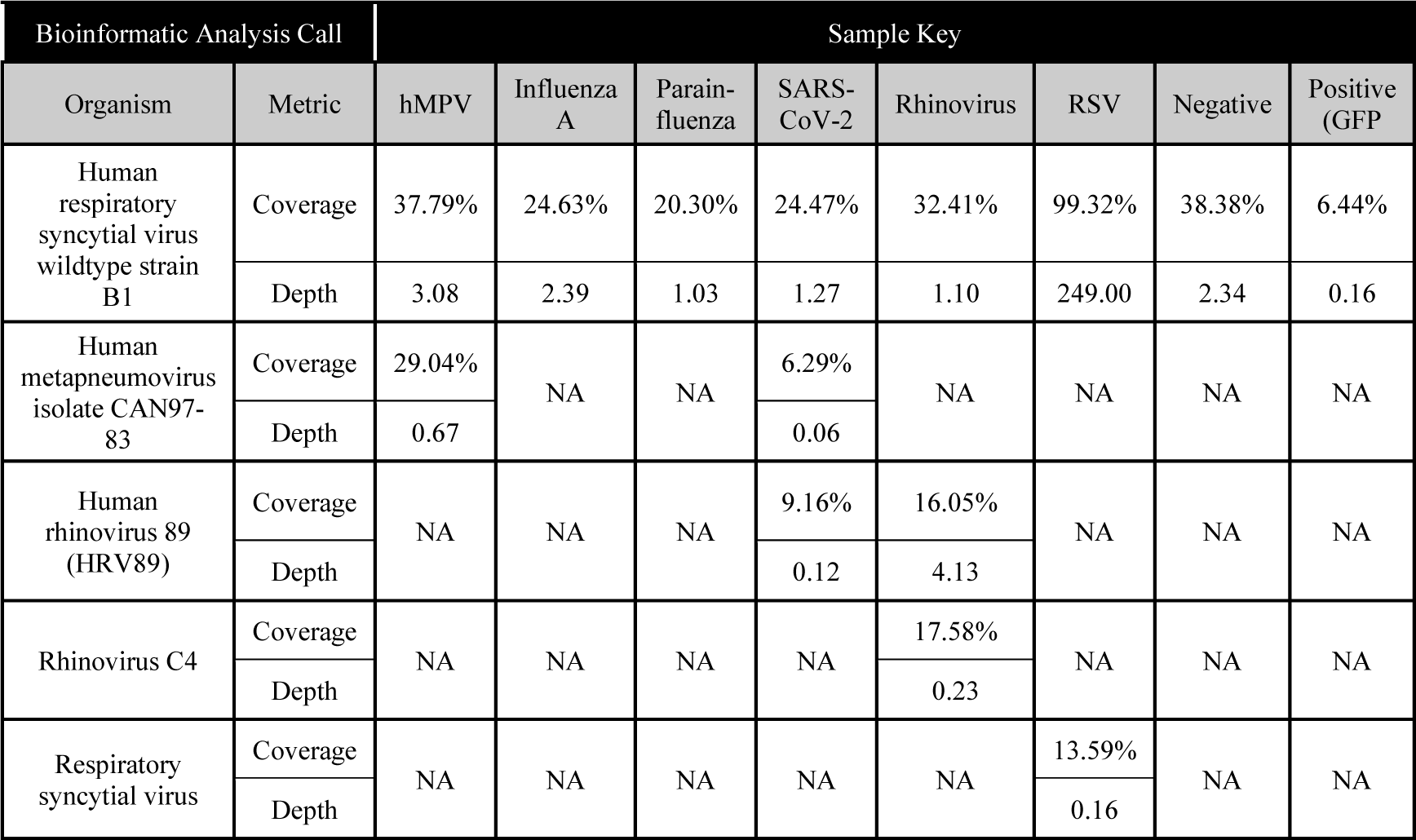
Clinical Remnant Sample Results with WTA assisted dscDNA method.

## 1 Discussion

### Potential Time from Sample Extraction to Sequencing Results

Reducing the time required for sample preparation and analysis for RNA or cDNA sequencing are important factors for public health utilization. The average time required to generate a single sample library (1-plex) using our unbiased workflow is 9 hrs from raw RNA to loading the MinION flowcell (Figure 9). Multiplexing (6-plex) on average increases each segment of the hands-on time by 15 min, while not affecting the incubation time except for End Repair and Ligation. To multiplex, samples must have a unique barcode ligated before continuing, resulting in a second ligation reaction and bead cleanup. While this workflow is longer than that of standard workday, there are safe stopping points can be found throughout our workflow as indicated in Figure 9 with an asterisk to break it over multiple days.

Another limiting factor in sequence analysis is the amount of time needed to generate the necessary sequence data to identify a potential pathogen within a sample. The process of a DNA sequencing run can take multiple days to complete on high throughput instruments. Sequencing runs used in this study were generally conducted for 22 hours to maximize sequence data capture. To evaluate whether the full 22 hours was needed for viral identification, we evaluated the viral genome coverage and read depth over time. Figure 10 shows key viral identification metrics from a contrived VEEV sample in plasma (1.95E03 GE/mL) following WGA in conjunction with the dscDNA method. Percent identity based on a consensus generated by the mapped reads and Blast+ indicated that the lowest identity for any covered portion was 97.25%. ‘Mapped Coverage’ measured by minmap2 rising above 90% by 4.25 hrs and maxing at 92.4%, while blast results showed coverage rising to 90% by 2.5 hrs and hitting a maximum of 95%. Based on the ‘Duty time’ record in Figure 8, these were achieved while the capacity of active sequencing pores was still greater than 50% of the flow cell and two-thirds from initiation of the run. This suggests the potential to shorten sequencing run times to < 6 hours depending on the nature of the sample.

### Implementing metagenomic assays in clinical laboratories

This study illustrates the potential of metagenomic analysis to public health labs. While the viruses used in this study could be detected by PCR approaches in a rapid and straightforward manner, targeted detection would require *a priori* knowledge of pathogen present or potentially expansive screening using viral detection panels. In the case of a novel, emerging, or uncommon viral pathogen, PCR may fail to identify the agent entirely. This is common in the case of viral sepsis, where a large proportion of cases result in a failure to identify the etiological agent [19]. Thus, it is critical to have a unbiased, metagenomic method capable of identifying novel, emerging, and rare viral threats compatible with widespread utilization in clinical and public health laboratories.

This approach does not utilize host RNA depletion. Not only did host RNA depletion not improve the relative proportion of viral sequences, but the method also used actually decreased the amount of viral signature in the sample. This may be due to the relatively low amount of RNA present in these samples and the potential for non-specific activity of the enzymatic digestion approach used. The clinical sample types used in this study were also likely responsible for the low host RNA background. VTM and plasma have lower host background than other sample types, such as whole blood. The application of a genome amplification step supports this observation in that, even following amplification, host background does not appear to overwhelm the viral genome signature within the sample, even at relatively low viral titers.

The method described herein does pose limitations in respect to widespread clinical laboratory implementation. While the molecular methods are not onerous, the workflow remains reasonably labor intensive. RNA extractions may be readily automated, and other methods have illustrated approaches for automation of nanopore sequencing library preparation [20,21]. Future development of automated for this method could streamline the workflow and allow higher-throughput analysis based on sample throughput needs. Further optimization could also shorten the amount of time required for DNA sequencing. While this study generally utilized 16-24h sequencing runs, it was clear that the majority of DNA sequencing was completed in the initial few hours following initialization. Further evaluation may reveal shorter sequencing durations that are sufficient to capture viral signature detection without risking false negatives.

This method utilizing molecular barcodes to permit multiplexing. This approach has been previously shown to carry some risk of misattribution of barcodes leading to the risk of false positives in one sample due to viral signatures in a different sample in the same sequencing run [5]. While we did observe evidence of barcode “cross-talk,” it was insufficient to lead to false positive detections in negative control samples, even when samples within the same sequencing run had extremely high titers. The potential for barcode cross-talk should be closely monitored in future studies, however, to identify conditions that lead to sufficient cross contamination to lead to potential false positive detection. It is likely that if the high accuracy (HAC) basecaller or alternative highly accurate basecallers can be used the assignment of barcode bin will also improve. Currently, the HAC cannot process reads at a rate to keep up with generation while the fast basecaller can. Crosstalk between barcodes is also likely to improve as ONT iterates barcode length and sequence and R10 flow-cell iteration with related chemistries.

Ultimately, before a metagenomic analysis approach can be utilized in clinical laboratories, a more extensive method validation is needed. This process will better establish key assay metrics including sensitivity, specificity, limits-of-detection, and reproducibility. Further analysis should also focus on clinical samples to identify any confounding issues associated with the use of mock clinical samples.

## Conclusions

We report an optimal workflow for the unbiased detection of RNA viruses in common clinical matrices. This workflow can support multiplex analysis of up to six samples with approximately 9 (1-plex) to 11 (6-plex) hours of sample preparation, price ranging from $750 (single-plex) to $1565 (six-plex), followed by a 10-24 hour sequencing run. The assay can detect viral genomes extracted from human plasma or VTM following a nasal/throat swab with as little as 1.95E03 GE/mL. Bioinformatics analysis is currently tailored toward target pathogens but is readily scalable to a pan-viral assessment to enable unbiased viral signature detection, including homology assessment to identify novel or emerging variants. This data supports the implementation of a sensitive, medium throughput assay capable of unbiased detection of clinical threats in public health laboratories.

## Data Availability

All data produced in the present study are available upon reasonable request to the authors. Metagenomic reads from samples from this study will depleted of human host sequences and will be submitted to the NCBI BioProject database before peer reviewed publication.

## Additional Requirements

For additional requirements for specific article types and further information please refer to “Article types” on every Frontiers journal page.

## Conflict of Interest

The authors declare that the research was conducted in the absence of any commercial or financial relationships that could be construed as a potential conflict of interest.

## Author Contributions

All authors have read and approved the manuscript. Conceptualization: ADK, KLT, MBS, FCH. Data curation: ADK, MBS, NCK. Laboratory analysis: ANS, NCK, MCM, ADK, LWA. Funding acquisition: ADK, FCH, KLT. Data analysis: ADK, ANS, MBS, NCK, KLT, FCH. Project administration: ADK, FCH. Writing – original draft: ADK, ANS, MBS, NCK, FCH. Writing – review & editing: ADK, ANS, MBS, NCK, LWA, MCM, KLT, FCH.

## Funding

This work was supported by the Centers for Disease Control and Prevention’s investments to combat antibiotic resistance under contract number 75D30121C12250.

## Acknowledgments

The authors would like to thank Leslie Parke for oversight and project management for this effort. The authors would also like to thank Jim Gibson for his assistance creating the workflow figure and Maddie Pont for her technical review. This work appears in the preprint.

## 1 Data Availability Statement

The datasets generated during this study for this study can be found in our NCBI BioProject [Update].

## List of abbreviations [Update]

DNA: Deoxyribonucleic Acid
cDNA: Copy DNA
dcDNA: Direct Copy DNA
DscDNA: Double Stranded cDNA
EID: Emerging infectious Diseases
GE: Genome Equivalents
HAC: High Accuracy Basecalling
HCoV: Human CoronaVirus
QC: Quality Control
qPCR: Quantitative Polymerase Chain Reaction
RNA: Ribonucleic Acid
dRNA: Direct RNA
rRNA: Ribosomal RNA
RT: Reverse Transcription
VEEV: Venezuelan Equine Encephalitis Virus
VTM: Viral Transport Media
WTA: Whole Transcriptome Amplification

## Notes

### Competing Interest Statement

The authors have declared no competing interest.

### Funding Statement

This research was funded by Centers for Disease Control and Prevention under award numbers 75D30121C12250. The views and conclusions contained herein are those of the authors and should not be interpreted as official policy or endorsements of the HHS, CDC, or the US Government.

### Author Declarations

The Human Protections Administrator (HPA) for Signature Science, LLC, as designated by the Federalwide Assurance (FWA) issued by the US Department of Health and Human Services, reviewed the certifications of Institutional Review Board (IRB) review and approval associated with the discarded, deidentified, commercially available human swab samples obtained from two vendors Discovery Life Sciences, BioIVT, and Precision BioSciences that were used in this research study. The Signature Science, LLC HPA found that these three vendors appropriately obtained and documented their IRB approval and donor consent for the collection of the human samples that were then obtained and used in this research project.

