## Supplemental Material for "Unbiased metagenomic detection of RNA viruses for rapid identification of viral pathogens in clinical samples"

Table S1. Sequencing Summary of Source Viral Material

| Method | DcDNA | DcDNA | dRNA | dRNA | Rapid | Rapid | dscDNA | dscDNA | dscDNA |
|---|---|---|---|---|---|---|---|---|---|
| Virus | SARS-CoV-2 | VEEV | SARS-CoV-2 | VEEV | SARS-CoV-2 | VEEV | SARS-CoV-2 | VEEV | 229E |
| Genome Size (Kb) | 29.8 | 11.4 | 29.8 | 11.4 | 29.8 | 11.4 | 29.8 | 11.4 | 27.3 |
| Input Volume (μL) | 7.5 | 7.5 | 9 | 9 | 13 | 13 | 13 | 13 | 12 |
| GE in total | 7.96E+06 | 5.03E+09 | 9.56E+06 | 6.03E+09 | 1.38E+07 | 8.72E+09 | 1.38E+07 | 8.72E+09 | 1.64E+09 |
| Mean Depth | 1.29 | 2339.85 | 0.13 | 18.94 | 35.44 | 298616 | 16.24 | 58657.1 | 1286.04 |
| Coverage (%, minimap2) | 39.13 | 100 | 5.29 | 70.72 | 62.81 | 100 | 93.38 | 100 | 100 |
| Coverage (%, blastn) | 40 | 100 | 5 | 73 | 63 | 100 | 88 | 100 | 100 |
| Identity of Coverage (%)[1] | 96.8 | 96.34 | 97.67 | 98.14 | 89.56 | 96.36 | 86.26 | 96.36 | 99.92 |
| Longest Alignment length (Kb) | 2.74 | 7.87 | 1.63 | 3.9 | 1.14 | 3.96 | 2.84 | 6.29 | 6.54 |
| Total Reads | 5,921 | 58,185 | 46,025 | 97,874 | 420,603 | 10,429,765 | 7,636 | 851,076 | 195,330 |
| Passing Quality Reads (%) | 78.03 | 74.01 | 47.97 | 48 | 77.38 | 85.97 | 76.82 | 85.82 | 86 |
| Mapped Passed Reads to Virus (%) | 1.3 | 82.13 | 0.03 | 0.47 | 2.48 | 88.18 | 10.01 | 92.77 | 13.06 |

1 - Average in cases of multiple mappings from discontinous coverages

Table S2. Sequencing Summary of Contrived Samples of Contrived Viral Samples Using dscDNA or Rapid Methods

| Method | DcDNA | DcDNA | dRNA | dRNA | dscDNA | dscDNA | dscDNA | dscDNA | dscDNA | dscDNA | Rapid | Rapid | Rapid | Rapid | Rapid | Rapid |
|---|---|---|---|---|---|---|---|---|---|---|---|---|---|---|---|---|
| Virus | VEEV | VEEV | VEEV | VEEV | VEEV | VEEV | 229E | 229E | None | None | VEEV | VEEV | 229E | 229E | **None** | **None** |
| Matrices | Plasma | Plasma | Plasma | Plasma | Plasma | Plasma | VTM | VTM | Plasma | VTM | Plasma | Plasma | VTM | VTM | Plasma | VTM |
| Replicate | 1 | 2 | 1 | 2 | 1 | 2 | 1 | 2 | 1 | 1 | 1 | 2 | 1 | 2 | 1 | 1 |
| Genome Size (Kb) | 11.4 | 11.4 | 11.4 | 11.4 | 11.4 | 11.4 | 27.3 | 27.3 | NA | NA | 11.4 | 11.4 | 27.3 | 27.3 | NA | NA |
| Input Volume (µL) | 7.5 | 7.5 | 9 | 9 | 12 | 12 | 12 | 12 | 12 | 12 | 12 | 12 | 12 | 12 | 12 | 12 |
| GE in total | 6.48E+07 | 6.48E+07 | 7.78E+07 | 7.78E+07 | 1.04E+08 | 1.04E+08 | 1.33E+09 | 1.33E+09 | 0 | 0 | 1.04E+08 | 1.04E+08 | 1.33E+09 | 1.33E+09 | 0 | 0 |
| GE/mL in Sample | 2.47E+09 | 2.47E+09 | 2.47E+09 | 2.47E+09 | 2.47E+09 | 2.47E+09 | 3.17E+10 | 3.17E+10 | 0 | 0 | 2.87E+08 | 2.87E+08 | 3.17E+10 | 3.17E+10 | 0 | 0 |
| Mean Depth | 24.5 | 22.2 | 0.17 | 0.06 | 937.1 | 2236.3 | 734.54 | 3461.83 | 0 | 0 | 15373 | 7265 | 9688.86 | 7301.84 | 0 | 0 |
| Coverage (%, minimap2) | 99.94 | 99.97 | 10.28 | 4.77 | 99.97 | 99.99 | 100 | 100 | 0 | 0 | 99.99 | 99.99 | 100 | 100 | 0 | 0 |
| Coverage (%, blastn) | 100 | 99 | 11 | 5 | 99 | 99 | 100 | 100 | 0 | 0 | 99 | 99 | 100 | 100 | 0 | 0 |
| Identity of Coverage (%)[1] | 96.34 | 96.26 | 99.52 | 100 | 96.34 | 96.33 | 99.91 | 99.91 | 0 | 0 | 96.36 | 96.35 | 99.93 | 99.93 | 0 | 0 |
| N50 | 788 | 957 | 199 | 184 | 1,209 | 1,221 | 1,535 | 1,962 | 658 | 1317 | 303 | 280 | 349 | 331 | 217 | 261 |
| Longest Alignment length (Kb) | 3.41 | 2.24 | 0.58 | 0.58 | 4.62 | 4.46 | 6.11 | 6.87 | 0 | 0 | 2.07 | 2.08 | 3.14 | 2.61 | 0 | 0 |
| Total Reads | 8,976 | 5,771 | 60,599 | 49,466 | 19,094 | 46,177 | 575,045 | 2,411,913 | 2047 | 361,490 | 1,498,299 | 911,560 | 3,313,337 | 2,788,740 | 26,785 | 1,536,316 |
| Passing Quality Reads (%) | 71.65 | 61.15 | 29.21 | 25.39 | 80.22 | 77.97 | 35.15 | 13.94 | 65.5 | 28.07 | 78.84 | 75 | 66.71 | 74.25 | 35.89 | 65.03 |
| Mapped Passed Reads to Virus (%) | 9.45 | 13.3 | 0.03 | 0.02 | 69.92 | 73 | 5.68 | 17.04 | 0 | 0 | 67.08 | 57.42 | 52.22 | 47.42 | 0 | 0 |
| Passing bp (%) | 60.61 | 51.91 | 28.81 | 23.35 | 79.99 | 76.73 | 63.75 | 62.63 | 65.5 | 28.07 | 69.36 | 60.75 | 66.88 | 73.04 | 35.89 | 60.26 |

1 - Average in cases of multiple mappings from discontinous coverages

Table S3. Sequencing Summary of Host Depletion from Contrived Viral Samples

| Method | dscDNA | dscDNA | dscDNA | Rapid | Rapid | Rapid |
|---|---|---|---|---|---|---|
| Sample | VEEV | VEEV | VEEV | VEEV | VEEV | VEEV |
| Treatment | None | None | Depleted | None | None | Depleted |
| Genome Size (Kb) | 11.4 | 11.4 | 11.4 | 11.4 | 11.4 | 11.4 |
| Input Volume from Extraction to method (μL)[1] | 12 | 12 | 11 | 12 | 12 | 11 |
| GE in total | 1.04E+08 | 1.04E+08 | 9.50E+07 | 1.04E+08 | 1.04E+08 | 9.50E+07 |
| GE/mL in Sample | 2.87E+08 | 2.87E+08 | 2.87E+08 | 2.87E+08 | 2.87E+08 | 2.87E+08 |
| Post-depletion GE in total | NA | NA | 1.60E+07 | NA | NA | 1.60E+07 |
| Mean Depth | 937.1 | 2236.3 | 0.73 | 15373 | 7265 | 0.03 |
| Coverage (%, minimap2) | 99.97 | 99.99 | 39.37 | 99.99 | 99.99 | 2.89 |
| Coverage (%, blastn) | 99 | 99 | 44 | 99 | 99 | 4 |
| Identity of Coverage (%)[2] | 96.34 | 96.33 | 99.34 | 96.36 | 96.35 | 100 |
| N50 | 1,209 | 1,221 | 381 | 303 | 280 | 226 |
| Longest Alignment length (Kb) | 4.62 | 4.46 | 0.19 | 2.07 | 2.08 | 0.9 |
| Total Reads | 19,094 | 46,177 | 1,095 | 1,498,299 | 911,560 | 5352 |
| Passing Quality Reads (%) | 80.22 | 77.97 | 28.95 | 78.84 | 75 | 20.59 |
| Mapped Passed Reads to Virus (%) | 69.92 | 73 | 5.36 | 67.08 | 57.42 | 0.18 |

1 - RNA input to depletion kit is less, all resulting depleted RNA was input for sequencing library

2 -Average in cases of multiple mappings from discontinouts coverage

Table S4. Sequencing Summary of Multiplex of dscDNA Method

| Method | dscDNA | dscDNA | dscDNA | dscDNA | dscDNA | dscDNA | dscDNA | dscDNA | dscDNA | dscDNA | dscDNA | dscDNA | dscDNA |
|---|---|---|---|---|---|---|---|---|---|---|---|---|---|
| Virus or Sample | VEEV | VEEV | VEEV | VEEV | VEEV | VEEV | VEEV | VEEV | VEEV | VEEV | A549 RNA | Plasma | Unclassified |
| GE in total | 1.04E+08 | 1.04E+08 | 1.04E+06 | 1.04E+06 | 1.04E+04 | 1.04E+04 | 1.04E+02 | 1.04E+02 | 1.04E+00 | 1.04E+00 | NA | NA | NA |
| GE/mL in Sample | 2.47E+09 | 2.47E+09 | 2.47E+07 | 2.47E+07 | 2.47E+05 | 2.47E+05 | 2.47E+03 | 2.47E+03 | 2.47E+01 | 2.47E+01 | NA | NA | NA |
| Replicate | 1 | 2 | 1 | 2 | 1 | 2 | 1 | 2 | 1 | 2 | NA | NA | NA |
| Barcode | 1 | 2 | 3 | 4 | 5 | 6 | 7 | 8 | 9 | 10 | 11 | 12 | Unclassified |
| Genome Size (Kb) | 11.4 | 11.4 | 11.4 | 11.4 | 11.4 | 11.4 | 11.4 | 11.4 | 11.4 | 11.4 | NA | NA | NA |
| Input Volume from Extraction (µL) | 12 | 12 | 12 | 12 | 12 | 12 | 12 | 12 | 12 | 12 | 12 | 12 | NA |
| Mean Depth | 1099.25 | 1354.03 | 8.08 | 10.47 | 2.29 | 1.59 | 1.13 | 1.21 | 1.71 | 1.73 | 0.43 | 1.39 | 104.46 |
| Coverage (%, minimap2) | 99.98 | 99.99 | 99.63 | 99.67 | 86.82 | 72.61 | 65.61 | 74.01 | 70.9 | 77.67 | 25.44 | 78.88 | 99.97 |
| Coverage (%, blastn) | 99 | 99 | 99 | 99 | 88 | 75 | 67 | 79 | 73 | 79 | 26 | 81 | 99 |
| Identity of Coverage (%)[1] | 96.35 | 96.33 | 96.19 | 96.32 | 97.99 | 98.28 | 98.97 | 99.12 | 98.01 | 98.19 | 99 | 98.66 | 96.3 |
| N50 | 1574 | 1410 | 781 | 598 | 571 | 329 | 747 | 470 | 390 | 695 | 1036 | 520 | 982 |
| Longest Alignment length (Kb) | 4.85 | 4.63 | 3.55 | 3.48 | 2.08 | 2.88 | 2.21 | 1.35 | 2.49 | 2.23 | 1.42 | 2.2 | 4.22 |
| Total Reads | 18486 | 26217 | 1901 | 3706 | 3818 | 7134 | 2455 | 4633 | 4381 | 3134 | 86795 | 14208 | 72,172 |
| Passing Quality Reads (%) | 80.36 | 81.36 | 69.59 | 60.55 | 54.45 | 54.74 | 69.25 | 63.44 | 53.71 | 69.11 | 89.13 | 84.13 | 20.32 |
| Mapped Passed Reads to Virus (%) | 67.22 | 66.19 | 5.06 | 4.1 | 1.06 | 0.44 | 0.65 | 0.54 | 0.68 | 0.92 | 0.01 | 0.13 | 7.28 |
| Passing bp (%) | 84.32 | 84.64 | 41.51 | 64.06 | 7.49 | 57.53 | 51.25 | 72.68 | 17.01 | 74.56 | 83.63 | 85.16 | 4.16 |

1 - Average in cases of multiple mappings from discontinous coverages

Table S5. Sequencing Summary of Multiplex of Rapid Method

| Method | Rapid | Rapid | Rapid | Rapid | Rapid | Rapid | Rapid | Rapid | Rapid | Rapid | Rapid | Rapid | Rapid |
|---|---|---|---|---|---|---|---|---|---|---|---|---|---|
| Virus or Sample | VEEV | VEEV | VEEV | VEEV | VEEV | VEEV | VEEV | VEEV | VEEV | VEEV | A549 RNA | Plasma | Unclassified |
| GE in total | 1.04E+08 | 1.04E+08 | 1.04E+06 | 1.04E+06 | 1.04E+04 | 1.04E+04 | 1.04E+02 | 1.04E+02 | 1.04E+00 | 1.04E+00 | NA | NA | NA |
| GE/mL in Sample | 2.47E+09 | 2.47E+09 | 2.47E+07 | 2.47E+07 | 2.47E+05 | 2.47E+05 | 2.47E+03 | 2.47E+03 | 2.47E+01 | 2.47E+01 | NA | NA | NA |
| Replicate | 1 | 2 | 1 | 2 | 1 | 2 | 1 | 2 | 1 | 2 | NA | NA | NA |
| Barcode | 1 | 2 | 3 | 4 | 5 | 6 | 7 | 8 | 9 | 10 | 11 | 12 | Unclassified |
| Genome Size (Kb) | 11.4 | 11.4 | 11.4 | 11.4 | 11.4 | 11.4 | 11.4 | 11.4 | 11.4 | 11.4 | NA | NA | NA |
| Input Volume from Extraction (µL) | 12 | 12 | 12 | 12 | 12 | 12 | 12 | 12 | 12 | 12 | 12 | 12 | NA |
| Mean Depth | 246.63 | 260.82 | 2.1 | 8.5 | 0.22 | 0.32 | 0.33 | 0.19 | 0.29 | 0.56 | 0.41 | 0.11 | 29.96 |
| Coverage (%, minimap2) | 99.87 | 99.98 | 85.99 | 99.74 | 21.95 | 28.28 | 27.29 | 14.76 | 24.69 | 45.72 | 25.92 | 9.76 | 99.76 |
| Coverage (%, blastn) | 99 | 99 | 87 | 99 | 24 | 34 | 32 | 17 | 28 | 47 | 29 | 13 | 99 |
| Identity of Coverage (%)[1] | 96.34 | 96.35 | 98.41 | 96.41 | 99.79 | 99.8 | 99.45 | 99.48 | 99.71 | 99.52 | 99.54 | 99.23 | 96.33 |
| N50 | 276 | 304 | 238 | 227 | 232 | 226 | 232 | 226 | 222 | 225 | 234 | 227 | 268 |
| Longest Alignment length (Kb) | 3.15 | 2.79 | 1.56 | 1.81 | 1.21 | 1.2 | 1.1 | 0.92 | 0.59 | 1.71 | 1.15 | 0.53 | 2.37 |
| Total Reads | 35907 | 32043 | 64791 | 28077 | 41866 | 30055 | 143979 | 46633 | 83828 | 15697 | 57872 | 26880 | 181789 |
| Passing Quality Reads (%) | 85.64 | 85.34 | 86.65 | 81.28 | 84.24 | 82.29 | 86.37 | 85.23 | 83.07 | 75.55 | 85.74 | 84.39 | 21.95 |
| Mapped Passed Reads to Virus (%) | 17.66 | 24.1 | 0.09 | 1.01 | 0.01 | 0.03 | 0.01 | 0.02 | 0.02 | 0.08 | 0.02 | 0.02 | 1.66 |
| Passing bp (%) | 86.72 | 86.38 | 86.63 | 81.12 | 83.45 | 81.39 | 84.18 | 84.61 | 82.16 | 75.36 | 87.23 | 84.2 | 10.2 |

1 - Average in cases of multiple mappings from discontinous coverages

Table S6. Sequencing Summary of Fongle Use

| Method | dscDNA | Rapid |
|---|---|---|
| Virus | VEEV | VEEV |
| GE in total | 1.04E+08 | 1.04E+08 |
| GE/mL in Sample | 2.47E+09 | 2.47E+09 |
| Genome Size (Kb) | 11.4 | 11.4 |
| Input Volume from Extraction (µL) | 12 | 12 |
| Mean Depth | 619.94 | 1.22 |
| Coverage (%, minimap2) | 99.99 | 70.37 |
| Coverage (%, blastn) | 99 | 74 |
| Identity of Coverage (%)[1] | 96.32 | 99.15 |
| N50 | 1434 | 208 |
| Longest Alignment length (Kb) | 4189 | 496 |
| Total Reads | 16724 | 4285 |
| Passing Quality Reads (%) | 57.21 | 20.65 |
| Mapped Passed Reads to Virus (%) | 66.71 | 9.27 |
| Passing bp (%) | 54.3 | 4.23 |

1 - Average in cases of multiple mappings from discontinous coverages

Table S7. Sequencing Summary of Whole Genome Amplifciation and dscDNA Method using VEEV

| Method | dscDNA | dscDNA | dscDNA | dscDNA | dscDNA | dscDNA | None | None | None | None | None | None | None |
|---|---|---|---|---|---|---|---|---|---|---|---|---|---|
| Virus or Sample | VEEV | VEEV | VEEV | VEEV | VEEV | VEEV | NA | NA | NA | NA | NA | NA | NA |
| GE in total | 8.21E+05 | 8.21E+05 | 8.21E+03 | 8.21E+03 | 8.21E+01 | 8.21E+01 | 0 | 0 | 0 | 0 | 0 | 0 | 0 |
| GE/mL in Sample | 1.95E+07 | 1.95E+07 | 1.95E+05 | 1.95E+05 | 1.95E+03 | 1.95E+03 | 0 | 0 | 0 | 0 | 0 | 0 | 0 |
| Replicate | 1 | 2 | 1 | 2 | 1 | 2 | 0 | 0 | 0 | 0 | 0 | 0 | 0 |
| Barcode | 1 | 2 | 3 | 4 | 5 | 6 | 7 | 8 | 9 | 10 | 11 | 12 | unclassified |
| Genome Size (Kb) | 11.4 | 11.4 | 11.4 | 11.4 | 11.4 | 11.4 | NA | NA | NA | NA | NA | NA | NA |
| Input Volume from Extraction (µL | 12 | 12 | 12 | 12 | 12 | 12 | NA | NA | NA | NA | NA | NA | NA |
| Mean Depth | 2954.7 | 3914.28 | 150.02 | 137.55 | 47.41 | 66.24 | 0 | 0 | 0 | 0.18 | 0 | 0 | 1071.17 |
| Coverage (%, minimap2) | 99.99 | 99.97 | 98.34 | 98.44 | 90.33 | 92.42 | 0 | 0 | 0 | 2.52 | 0 | 0 | 99.98 |
| Coverage (%, blastn) | 99 | 99 | 99 | 99 | 94 | 95 | 0 | 0 | 0 | 10 | 0 | 0 | 99 |
| Identity of Coverage (%)[1] | 96.34 | 96.37 | 96.29 | 96.33 | 96.94 | 97.7 | 0 | 0 | 0 | 97.57 | 0 | 0 | 96.34 |
| N50 | 3376 | 2940 | 2734 | 3476 | 3483 | 3496 | 6202 | 0 | 3145 | 4028 | 4225 | 1642 | 3813 |
| Longest Alignment length (Kb | 3.19 | 2.96 | 1.84 | 3.15 | 1.84 | 2.86 | 0 | 0 | 0 | 0.29 | 0 | 0 | 2.13 |
| Total Reads for a Barcode | 98649 | 139714 | 136040 | 108623 | 82638 | 120026 | 13 | 0 | 22 | 9 | 15 | 32 | 237016 |
| Percent of Total Reads in Run | 10.69 | 15.14 | 14.74 | 11.77 | 8.96 | 13.01 | 0 | 0 | 0 | 0 | 0 | 0 | 25.68 |
| Passing Quality Reads (%) | 68.51 | 68.4 | 69.74 | 68.21 | 71.28 | 68.91 | 15.38 | 0 | 13.64 | 22.22 | 6.67 | 9.38 | 26.84 |
| Mapped Passed Reads to Virus (%) | 25.89 | 26.83 | 0.96 | 1.01 | 0.48 | 0.51 | 0 | 0 | 0 | 50 | 0 | 0 | 10.21 |
| Passing bp (%) | 66.86 | 66.2 | 67.02 | 65.28 | 68.15 | 66.83 | 22.72 | 0 | 16.68 | 23.07 | 18.54 | 4.13 | 31.48 |

1 - Average in cases of multiple mappings from discontinous coverages

Table S8. Sequencing Summary of Whole Genome Amplification and Rapid Method using VEEV

| Method | Rapid | Rapid | Rapid | Rapid | Rapid | Rapid | None | None | None | None | None | None | None |
|---|---|---|---|---|---|---|---|---|---|---|---|---|---|
| Virus or Sample | VEEV | VEEV | VEEV | VEEV | VEEV | VEEV | NA | NA | NA | NA | NA | NA | NA |
| GE in total | 8.21E+05 | 8.21E+05 | 8.21E+03 | 8.21E+03 | 8.21E+01 | 8.21E+01 | 0 | 0 | 0 | 0 | 0 | 0 | 0 |
| GE/mL in Sample | 1.95E+07 | 1.95E+07 | 1.95E+05 | 1.95E+05 | 1.95E+03 | 1.95E+03 | 0 | 0 | 0 | 0 | 0 | 0 | 0 |
| Replicate | 1 | 2 | 1 | 2 | 1 | 2 | 0 | 0 | 0 | 0 | 0 | 0 | 0 |
| Barcode | 1 | 2 | 3 | 4 | 5 | 6 | 7 | 8 | 9 | 10 | 11 | 12 | unclassified |
| Genome Size (Kb) | 11.4 | 11.4 | 11.4 | 11.4 | 11.4 | 11.4 | NA | NA | NA | NA | NA | NA | NA |
| Input Volume from Extraction (µL) | 12 | 12 | 12 | 12 | 12 | 12 | NA | NA | NA | NA | NA | NA | NA |
| Mean Depth | 5056.2 | 5176.32 | 100.84 | 114.58 | 5.31 | 3.62 | 0.29 | 0.04 | 0.12 | 0 | 0 | 0.09 | 606.77 |
| Coverage (%, minimap2) | 99.59 | 99.27 | 97.91 | 97.98 | 62.48 | 71.49 | 9.38 | 4.24 | 9.92 | 0 | 0 | 3.87 | 99.87 |
| Coverage (%, blastn) | 99 | 99 | 99 | 99 | 67 | 78 | 14 | 5 | 20 | 0 | 0 | 7 | 99 |
| Identity of Coverage (%)[1] | 96.34 | 96.34 | 96.35 | 96.25 | 96.97 | 96.82 | 97.35 | 99.59 | 98.63 | 0 | 0 | 99.1 | 96.37 |
| N50 | 2578 | 2658 | 2572 | 2661 | 2464 | 2529 | 6965 | 772 | 1819 | 357 | 1375 | 1695 | 2599 |
| Longest Alignment length (Kb) | 2.71 | 2.62 | 2.1 | 1.93 | 1.75 | 1.89 | 0.59 | 0.49 | 0.77 | 0 | 0 | 0.44 | 2.17 |
| Total Reads for a Barcode | 188566 | 185313 | 156451 | 162121 | 213392 | 133602 | 24 | 18 | 18 | 2 | 8 | 6 | 215288 |
| Percent of Total Reads in Run | 15.03 | 14.77 | 12.47 | 12.92 | 17.01 | 10.65 | 0 | 0 | 0 | 0 | 0 | 0 | 17.16 |
| Passing Quality Reads (%) | 90.06 | 89.37 | 89.74 | 89.05 | 89.9 | 89.93 | 66.67 | 83.33 | 72.22 | 50 | 50 | 66.67 | 32.88 |
| Mapped Passed Reads to Virus (%) | 25.43 | 25.98 | 0.58 | 0.6 | 0.02 | 0.03 | 12.5 | 6.67 | 15.38 | 0 | 0 | 25 | 6.85 |
| Passing bp (%) | 88.52 | 88.1 | 88.27 | 87.4 | 88.93 | 89.2 | 81.52 | 89.14 | 73.93 | 18.35 | 63.88 | 62.35 | 37.87 |

1 - Average in cases of multiple mappings from discontinous coverages

Table S9. Sequencing Summary of Whole Genome Amplification and dscDNA Method using HCoV 229E

| Method | **None** | **None** | **None** | **None** | **None** | **None** | dscDNA | dscDNA | dscDNA | dscDNA | dscDNA | dscDNA | None |
|---|---|---|---|---|---|---|---|---|---|---|---|---|---|
| Virus or Sample | NA | NA | NA | NA | NA | NA | 229E | 229E | 229E | 229E | 229E | 229E | NA |
| GE in total | 0 | 0 | 0 | 0 | 0 | 0 | 2.29E+07 | 2.29E+07 | 2.29E+06 | 2.29E+06 | 2.29E+05 | 2.29E+05 | 0 |
| GE/mL in Sample | 0 | 0 | 0 | 0 | 0 | 0 | 5.43E+08 | 5.43E+08 | 5.43E+07 | 5.43E+07 | 5.43E+06 | 5.43E+06 | 0 |
| Replicate | 0 | 0 | 0 | 0 | 0 | 0 | 1 | 2 | 1 | 2 | 1 | 2 | 0 |
| Barcode | 1 | 2 | 3 | 4 | 5 | 6 | 7 | 8 | 9 | 10 | 11 | 12 | unclassified |
| Genome Size (Kb) | 0 | 0 | 0 | 0 | 0 | 0 | 27.3 | 27.3 | 27.3 | 27.3 | 27.3 | 27.3 | 0 |
| Input Volume from Extraction (µL) | 12 | 12 | 12 | 12 | 12 | 12 | 12 | 12 | 12 | 12 | 12 | 12 | 12 |
| Mean Depth | 0.08 | 0 | 0 | 0.03 | 0.09 | 0.12 | 3566.62 | 3050.51 | 1572.42 | 1562.44 | 2059.74 | 1612.87 | 2010.63 |
| Coverage (%, minimap2) | 5.64 | 0 | 0 | 0.81 | 3.27 | 4.93 | 99.98 | 100 | 99.94 | 99.92 | 99.86 | 99.95 | 99.98 |
| Coverage (%, blastn) | 7 | 0 | 0 | 6 | 4 | 8 | 100 | 100 | 100 | 100 | 100 | 100 | 100 |
| Identity of Coverage (%)[1] | 99.95 | 0 | 0 | 99.55 | 99.91 | 99.58 | 99.89 | 99.91 | 99.91 | 99.92 | 99.89 | 99.89 | 99.92 |
| N50 | 4521 | 2301 | 4732 | 1638 | 3518 | 1765 | 3164 | 3867 | 3434 | 3255 | 3305 | 3097 | 3757 |
| Longest Alignment length (Kb) | 986 | 0 | 0 | 221 | 574 | 554 | 3419 | 3193 | 3216 | 314 | 2822 | 2340 | 3208 |
| Total Reads for a Barcode | 26 | 54 | 38 | 22 | 19 | 31 | 159507 | 134289 | 150470 | 152969 | 175653 | 135563 | 309168 |
| Percent of Total Reads in Run | 0 | 0 | 0 | 0 | 0 | 0 | 13.1 | 11.03 | 12.36 | 12.56 | 14.42 | 11.13 | 25.39 |
| Passing Quality Reads (%) | 34.62 | 14.81 | 13.16 | 18.18 | 21.05 | 22.58 | 83.53 | 78.13 | 80.41 | 81 | 79.07 | 80.52 | 33.98 |
| Mapped Passed Reads to Virus (%) | 22.22 | 0 | 0 | 25 | 50 | 42.86 | 36.5 | 34.49 | 18.15 | 18.32 | 23.24 | 23.6 | 26.19 |
| Passing bp (%) | 33.02 | 26.19 | 18.97 | 25.11 | 25.47 | 17.27 | 83.03 | 81.77 | 80.3 | 80.22 | 78.02 | 79.37 | 40.71 |

1 - Average in cases of multiple mappings from discontinous coverages

Table S10. Coverage of Genomes from Clinical Remnant Samples

| Bioinformatic Anaylsis Call In Run | | Sample Key | | | | | | | |
|---|---|---|---|---|---|---|---|---|---|
| | | hMPV | Influenza A | Parainfluenza IV | SARS-CoV-2 | Rhinovirus | RSV | EB (negative) | GFP |
| Accession | Organism | barcode17 | barcode18 | barcode19 | barcode20 | barcode21 | barcode22 | barcode23 | barcode24 |
| AF013254.1 | Human respiratory syncytial virus wildtype strain B1 | 37.79% | 24.63% | 20.30% | 24.47% | 32.41% | 99.32% | 38.38% | 6.44% |
| AY297749.1 | Human metapneumovirus isolate CAN97-83 | 29.04% | NA | NA | 6.29% | NA | NA | NA | NA |
| AF304460.1 | Human coronavirus 229E | 2.20% | 4.47% | 3.74% | 3.35% | NA | 6.18% | 10.31% | 2.31% |
| U05876.1 | coxsackievirus A16 G-10 | NA | 2.17% | NA | NA | NA | NA | NA | NA |
| JN831356.1 | Canine picornavirus strain 325F | NA | NA | 1.70% | NA | 2.28% | 1.66% | 1.69% | NA |
| A10937.1 | Human rhinovirus 89 (HRV89) | NA | NA | NA | 9.16% | 16.05% | NA | NA | NA |
| CU928144.1 | Escherichia fergusonii ATCC 35469 plasmid pEFER | NA | NA | NA | 1.13% | NA | NA | 2.20% | NA |
| D00820.1 | Human enterovirus 70 genomic RNA strain: J670/71 | NA | NA | NA | NA | 12.42% | NA | NA | NA |
| AF326766.2 | Enterovirus J1 strain 1631 isolate Simian enterovirus SV6 | NA | NA | NA | NA | 6.01% | 2.44% | NA | NA |
| V01149.1 | Human poliovirus 1 Mahoney | NA | NA | NA | NA | 5.58% | NA | NA | NA |
| HQ595344.1 | Bat picornavirus 3 strain TLC5F | NA | NA | NA | NA | 2.13% | NA | NA | NA |
| EF582385.1 | Rhinovirus C4 | NA | NA | NA | NA | 17.58% | NA | NA | NA |
| U39661.1 | Respiratory syncytial virus | NA | NA | NA | NA | NA | 13.59% | NA | NA |
| J02459.1 | Escherichia phage Lambda | NA | NA | NA | NA | NA | NA | 2.74% | NA |
| CP046033.1 | Salmonella sp. HNK130 chromosome | NA | NA | NA | NA | NA | NA | 1.65% | NA |
| U00096.3 | Escherichia coli str. K-12 substr. MG1655 | NA | NA | NA | NA | NA | NA | 1.58% | NA |
| CP014768.1 | Shigella sp. PAMC 28760 chromosome | NA | NA | NA | NA | NA | NA | 1.00% | NA |
| AE017283.1 | Propionibacterium acnes KPA171202 | NA | NA | NA | NA | NA | NA | 1.30% | NA |
| AP018683.1 | Vibrio casei DSM 22364 plasmid 2 DNA | NA | NA | NA | NA | NA | NA | 1.24% | NA |

Matching highlighting beteween Bioinformatics Analysis Call In Run and the Sample Key indiciate potetnial congruency

Table S11. Mean Depth of Genomes from Clinical Remnant Samples

| Bioinformatic Anaylsis Call In Run | | Sample Key | | | | | | | |
|---|---|---|---|---|---|---|---|---|---|
| | | hMPV | Influenza A | Parainfluenza IV | SARS-CoV-2 | Rhinovirus | RSV | EB (negative) | GFP |
| Accession | Organism | barcode17 | barcode18 | barcode19 | barcode20 | barcode21 | barcode22 | barcode23 | barcode24 |
| AF013254.1 | Human respiratory syncytial virus wildtype strain B1 | 3.08841 | 2.38844 | 1.02516 | 1.26851 | 1.09655 | 248.998 | 2.3357 | 0.164532 |
| AY297749.1 | Human metapneumovirus isolate CAN97-83 | 0.672516 | NA | NA | NA | NA | NA | NA | NA |
| CP032084.1 | Achromobacter sp. B7 chromosome | 0.313083 | 0.829797 | 0.378324 | NA | NA | 0.681702 | NA | NA |
| AE017283.1 | Propionibacterium acnes KPA171202 | 0.294837 | 0.190276 | 0.122335 | 0.161913 | NA | 0.146996 | 0.2368 | NA |
| CP002287.1 | Achromobacter xylosoxidans A8 | 0.244979 | 0.636773 | 0.268886 | NA | NA | 0.508442 | NA | NA |
| CP018101.1 | Delftia sp. HK171 chromosome | 0.196228 | 0.382438 | 0.132444 | NA | NA | 0.271848 | NA | NA |
| CP002735.1 | Delftia sp. Cs1-4 | 0.112842 | 0.234211 | NA | NA | NA | 0.173979 | NA | NA |
| AF304460.1 | Human coronavirus 229E | NA | 5.3764 | 6.73017 | NA | NA | 5.38401 | 19.8899 | NA |
| CP026973.1 | Achromobacter insolitus strain FDAARGOS_88 chromosome | NA | 0.222466 | NA | NA | NA | 0.17009 | NA | NA |
| CP041204.1 | Rhizobium sp. NIBRBAC000502774 chromosome | NA | 0.194988 | NA | NA | NA | 0.152877 | 0.163118 | NA |
| JN831356.1 | Canine picornavirus strain 325F | NA | NA | 0.107826 | NA | 8.47811 | NA | NA | NA |
| A10937.1 | Human rhinovirus 89 (HRV89) | NA | NA | NA | 0.122204 | 4.13255 | NA | NA | NA |
| D00820.1 | Human enterovirus 70 genomic RNA strain: J670/71 | NA | NA | NA | NA | 6.71218 | NA | NA | NA |
| AF326766.2 | Enterovirus J1 strain 1631 isolate Simian enterovirus SV6 | NA | NA | NA | NA | 1.28676 | NA | NA | NA |
| V01149.1 | Human poliovirus 1 Mahoney | NA | NA | NA | NA | 0.48414 | NA | NA | NA |
| HQ595344.1 | Bat picornavirus 3 strain TLC5F | NA | NA | NA | NA | 0.312169 | NA | NA | NA |
| EF582385.1 | Rhinovirus C4 | NA | NA | NA | NA | 0.233554 | NA | NA | NA |
| U39661.1 | Respiratory syncytial virus | NA | NA | NA | NA | NA | 0.160292 | NA | NA |

Matching highlighting beteween Bioinformatics Analysis Call In Run and the Sample Key indiciate potetnial congruency
